# Development, evaluation, and validation of machine learning models for COVID-19 detection based on routine blood tests

**DOI:** 10.1101/2020.10.02.20205070

**Authors:** Cabitza Federico, Campagner Andrea, Ferrari Davide, Di Resta Chiara, Ceriotti Daniele, Sabetta Eleonora, Colombini Alessandra, De Vecchi Elena, Banfi Giuseppe, Locatelli Massimo, Carobene Anna

## Abstract

**Background:** The rRT-PCR test, the current gold standard for the detection of coronavirus disease (COVID-19), presents with known shortcomings, such as long turnaround time, potential shortage of reagents, false-negative rates around 15–20%, and expensive equipment. The hematochemical values of routine blood exams could represent a faster and less expensive alternative.

**Methods:** Three different training data set of hematochemical values from 1,624 patients (52% COVID-19 positive), admitted at San Raphael Hospital (OSR) from February to May 2020, were used for developing machine learning (ML) models: the complete OSR dataset (72 features: complete blood count (CBC), biochemical, coagulation, hemogasanalysis and CO-Oxymetry values, age, sex and specific symptoms at triage) and two sub-datasets (COVID-specific and CBC dataset, 32 and 21 features respectively). 58 cases (50% COVID-19 positive) from another hospital, and 54 negative patients collected in 2018 at OSR, were used for internal-external and external validation.

**Results:** We developed five ML models: for the complete OSR dataset, the area under the receiver operating characteristic curve (AUC) for the algorithms ranged from 0.83 to 0.90; for the COVID-specific dataset from 0.83 to 0.87; and for the CBC dataset from 0.74 to 0.86. The validations also achieved good results: respectively, AUC from 0.75 to 0.78; and specificity from 0.92 to 0.96.

**Conclusions:** ML can be applied to blood tests as both an adjunct and alternative method to rRT-PCR for the fast and cost-effective identification of COVID-19-positive patients. This is especially useful in developing countries, or in countries facing an increase in contagions.

## Introduction

To date, at eight months post-outbreak, the coronavirus disease (COVID-19) caused by the SARS-CoV-2 coronavirus has infected more than 20 million people and has resulted in approximately one million deaths worldwide. To manage this unprecedented pandemic emergency, the early identification of patients and of infectious people is extremely important due to the fact that this disease, unlike others caused by coronaviruses (e.g. SARS, MERS), can coexist in a host organism without causing any symptoms, or it can produce very mild and non-characteristic symptoms in—nevertheless—infectious subjects (1). To identify SARS-CoV-2 infections, the instrument of choice, or the gold standard, is the molecular test performed using the reverse polymerase chain reaction (PCR) or the reverse transcriptase–PCR (RT-PCR) technique. However, the execution of the test is time-consuming (at no less than 4–5 hours under optimal conditions), requires the use of special equipment and reagents, the involvement of specialized and trained personnel for the collection of the samples, and relies on the proper genetic conservation of the RNA sequences that are selected for annealing the primers (2). In addition, for these pre-analytical vulnerabilities (3), the RT-PCR test’s accuracy, and especially its sensitivity (i.e. its ability to avoid false negatives), is far from ideal. A recently published article in the *New England Journal of Medicine* suggests that a reasonable estimate for the sensitivity of this test is 70% (4).

To improve our diagnostic capabilities, in order to contain the spread of the pandemic, the data science community has proposed several machine learning (ML) models, recently reviewed in (5). Most of these models are based on computed tomography (CT) scans or chest x-rays (5–9). Despite the reported promising results, some concerns have been raised regarding these and other works, especially in regard to solutions based on chest x-rays, which have been associated with high rates of false-negative results (10). On the other hand, solutions based on CT imaging, although accurate, are affected by the characteristics of this modality: CTs are costly, time-consuming, and require specialized equipment; thus, approaches based on this imaging technique cannot reasonably be applied for screening exams. Although various clinical studies (11–13) have highlighted how blood test-based diagnostics might provide an effective and low-cost alternative for the early detection of COVID-19 cases, relatively few ML models have been applied to hematological parameters (14–18).

To overcome the above limitations, and following a successful feasibility study performed on a smaller dataset (19), we developed different classification models by applying ML techniques to blood-test results that are generally available in clinical practice within minutes (under emergency conditions, at even less than 15 minutes) and are only a fraction of the cost of the RT-PCR test and CT imaging (i.e. a few euros). As we will show, routine blood tests can be exploited by our method to diagnose COVID-19 patients in low-resource settings, in particular, where there is a shortage of RT-PCR reagents, such as during a pandemic peak. On the other hand, the developed method can also be used as a complement to the RT-PCR test in order to increase the sensitivity of the latter or to provide its interpreters a sort of pre-test probability to compute NPV and PPV. Furthermore, the rapid blood-test results could be a valuable—although non-conclusive—indication for the early identification of COVID-19 patients, resulting in their better management/isolation while waiting for the gold standard results.

## Methods

In this section, we describe the datasets and statistical methods used to train and validate the ML models. The reporting follows the TRIPOD Guideline for Model Development and Validation (20). The study protocol (BIGDATA-COVID19) was approved by the Institutional Ethical Review Board in agreement with the World Medical Association Declaration of Helsinki.

### Data Description

#### OSR dataset

The main dataset used for this study (the OSR dataset) consisted of routine blood-test results performed on 1,925 patients on admission to the ED at the San Raffaele Hospital (OSR) from February 19, 2020, to May 31, 2020. In order to control for potentially confounding pathologies and other sources of bias, such as insufficient data availability, in ML development, 301 (15.6%) patients, admitted between February and April, were excluded from further analysis. All patients admitted during May 2020, on the other hand, were considered for the study, to have a balanced number of patients also from the late portions of the time frame considered.

For each case, COVID-19 positivity was determined based on the result of the molecular test for SARS-CoV-2 performed by RT-PCR on nasopharyngeal swabs. On a set of 165 uncertain cases, we also used the result of chest radiography and x-rays to improve over the sensitivity of the RT-PCR test (21–24). Uncertain cases were identified through two different methods: either patients who resulted positive within 72 hours after a first negative test and were admitted as inpatients despite this test result; or patients who, despite having a negative test, had an hematochemical profile more similar to positive patients, as determined through multi-variate clustering based on a set of COVID-19 characteristic biomarkers (12) (AST, lymphocytes, calcium, LDH, PCR, WBC, XDP, Fibrinogen). Of the 165 uncertain cases, only 52 of them have been considered as positive after comparison with the radiologic gold standard, while the remaining 113 were considered as negative (having a double negative test from both the RT-PCR and the radiologic gold standard): this results in an estimate of 93% sensitivity of the RT-PCR with respect to the composite ground truth.

Therefore, the OSR dataset consisted of a total of 1,624 cases: 786 of them received a positive diagnosis (48%) and 838 were negative cases (52%).

As covariate features, for each case, the patient’s age and gender, the presence of COVID-19 related symptomatology at admission (dyspnea, pneumonia, pyrexia, sore throat, influenza, cough, pharyngitis, bronchitis, generalized illness), and a set of 69 hematochemical values from laboratory tests were considered. The list of the analytes and instruments are reported in Table 1. The laboratory blood tests were performed according to the International Federation of Clinical Chemistry and Laboratory Medicine (IFCC) recommendations (25).

**Table 1.**
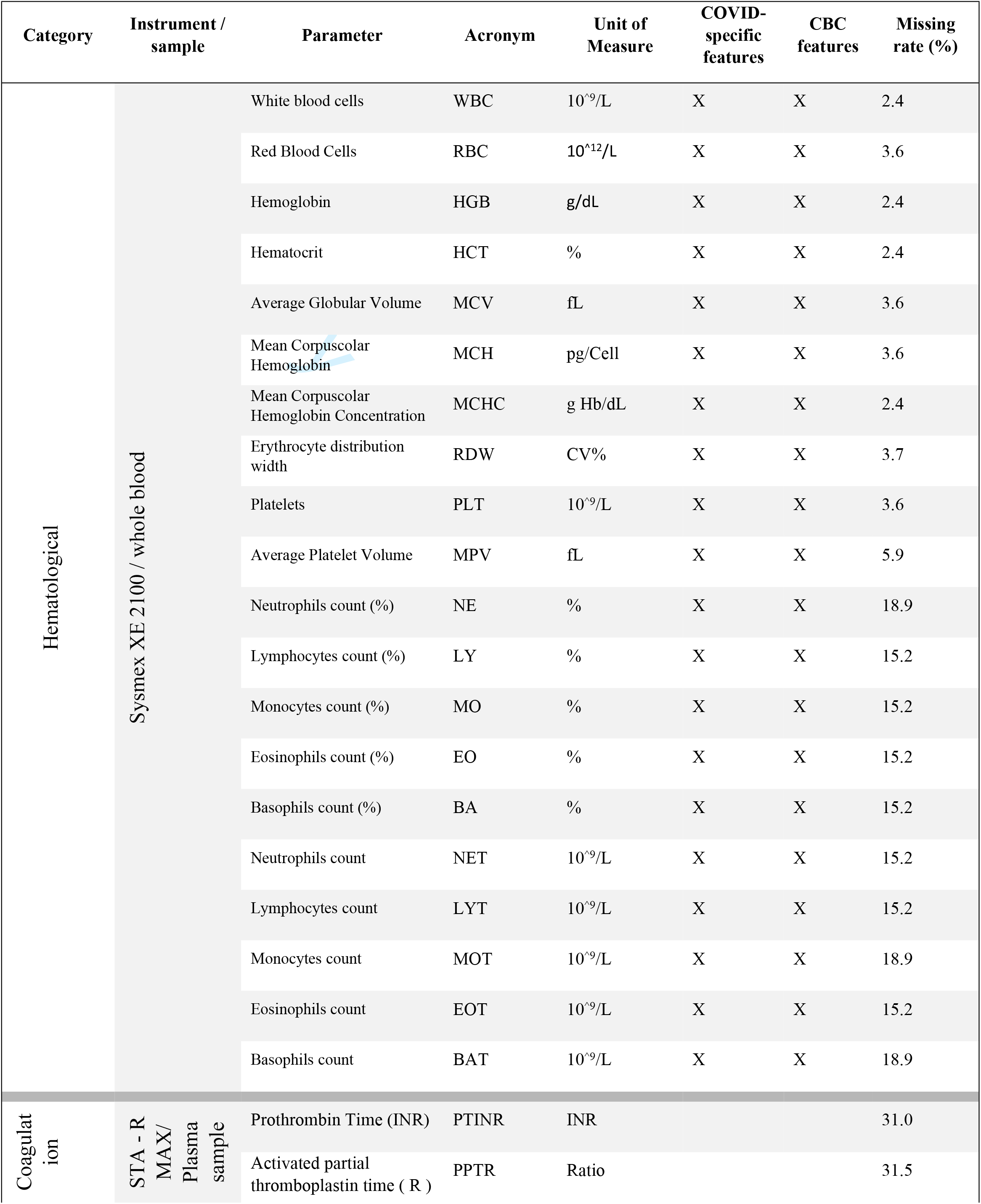
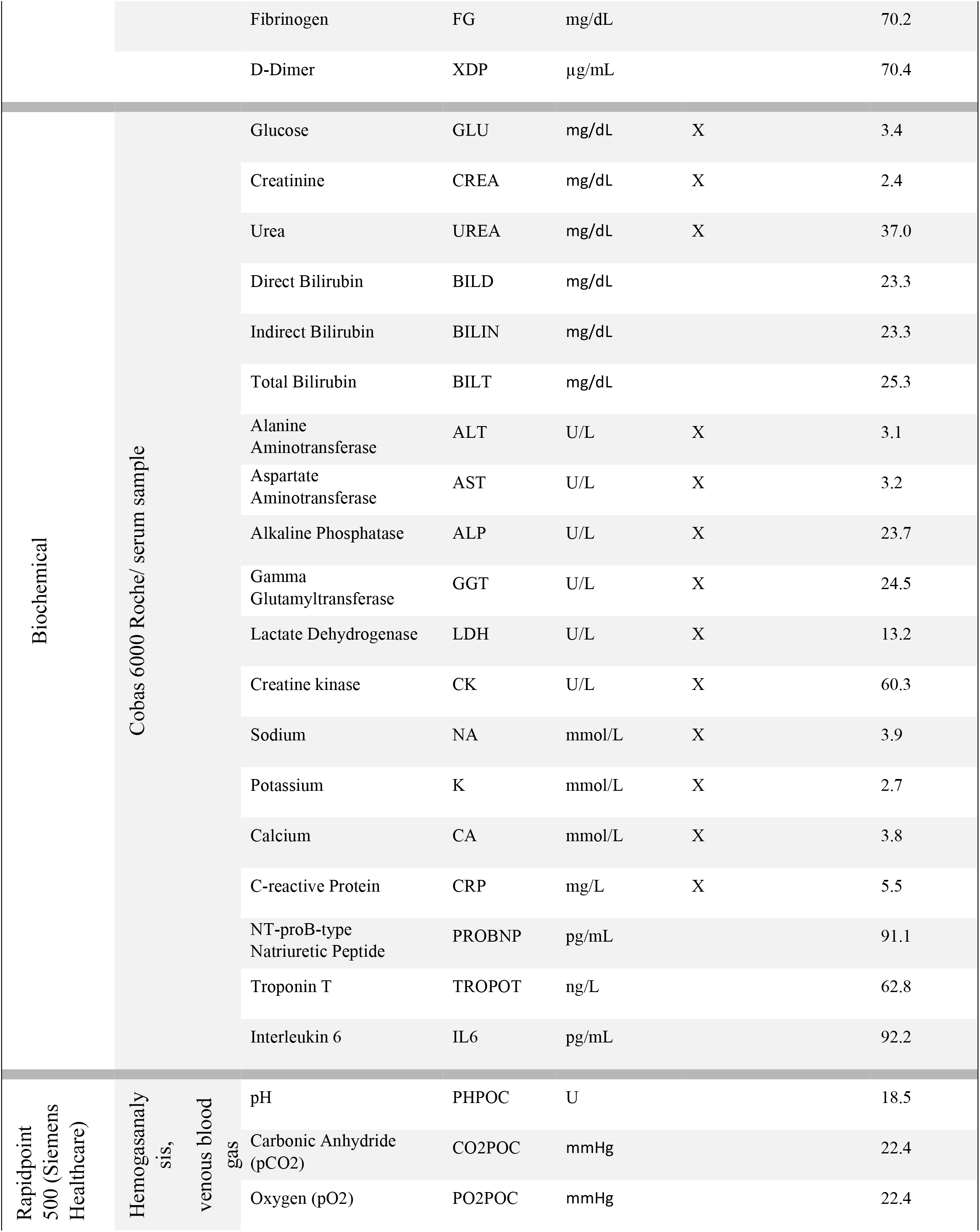
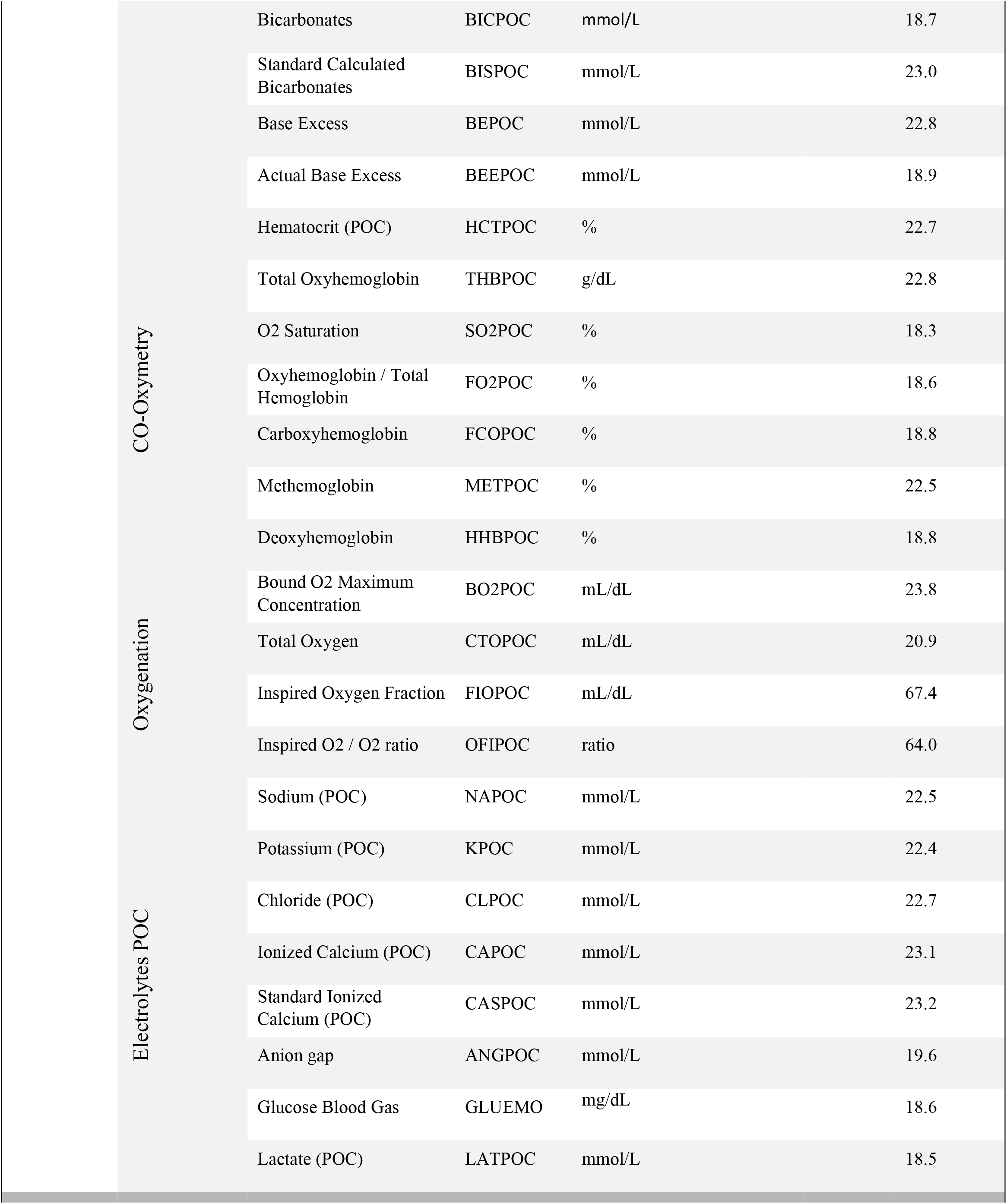
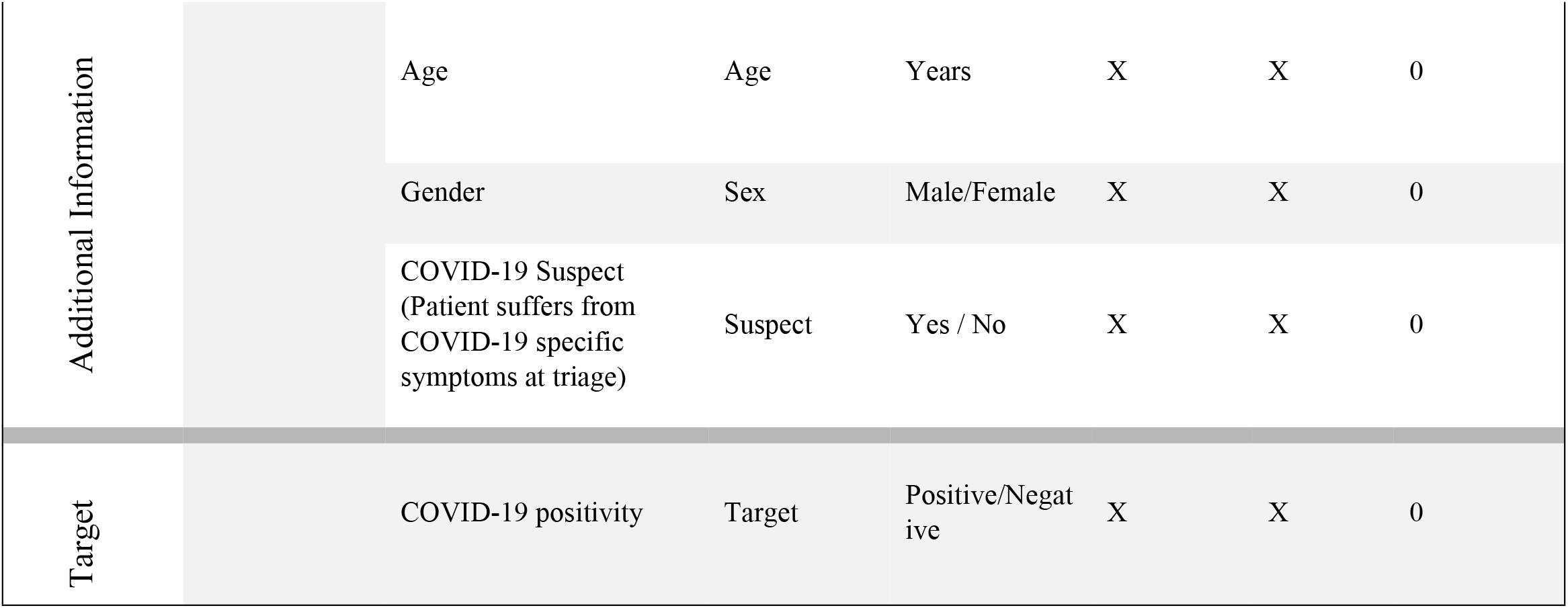
Complete list of the analyzed features in the *OSR dataset*.

The demographic and clinical characteristics of the two different groups of COVID-19 patients are summarized in Figure 1 and Figure 2.

**Figure 1.**
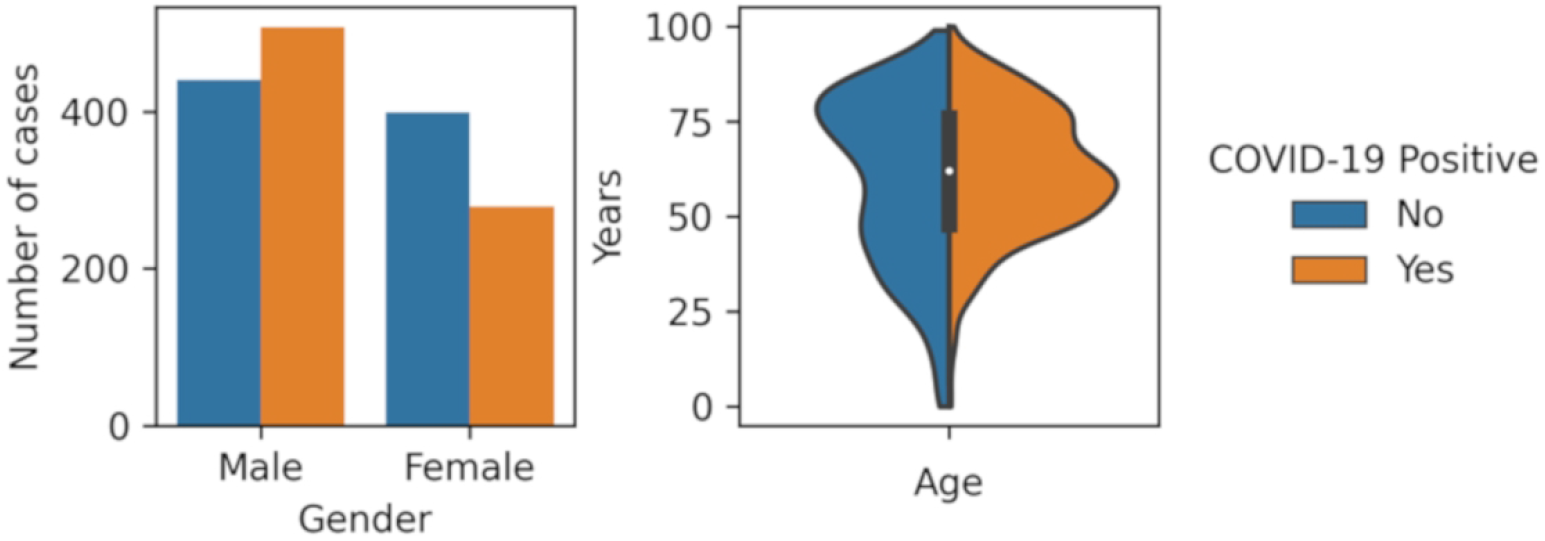
Demographic feature (gender and age) distributions for positive and negative cases. The blue and orange areas correspond to negative and positive cases, respectively

**Figure 2.**
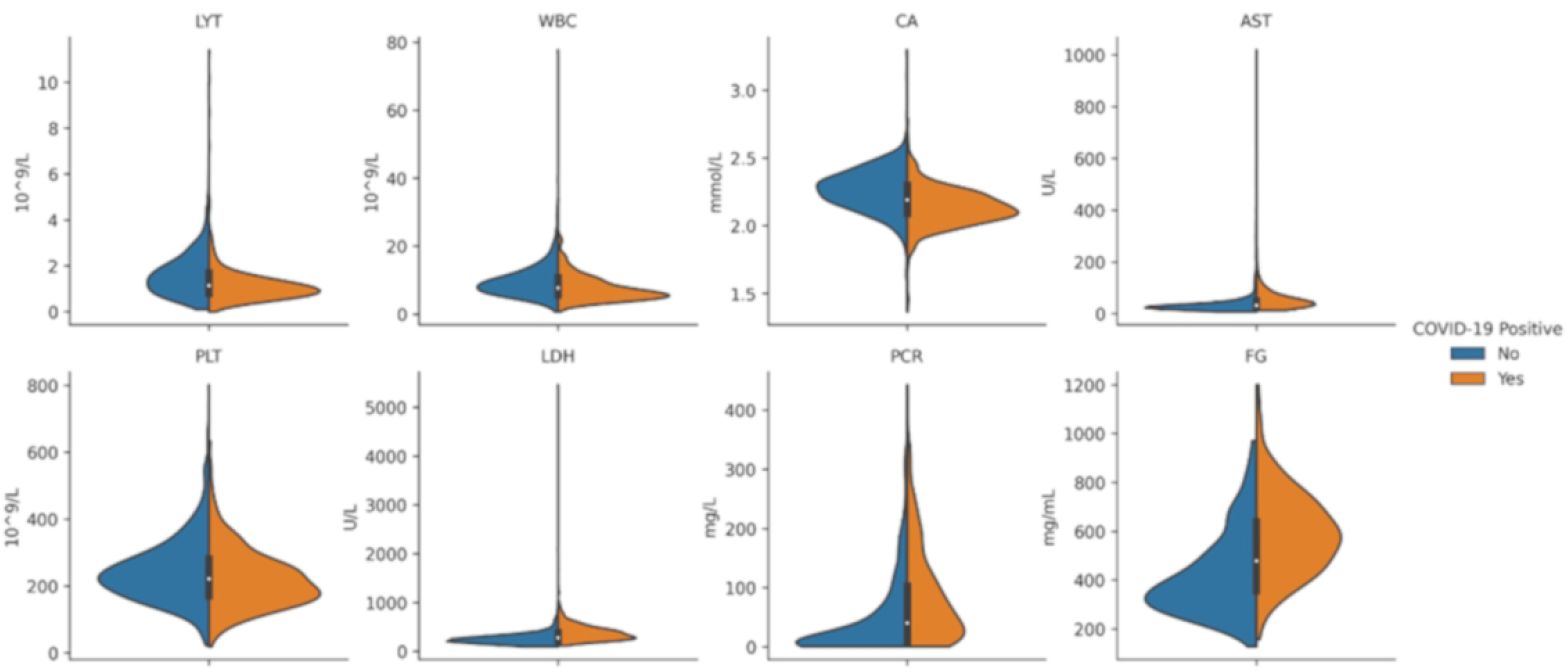
Violin plots depicting the distributions of eight relevant features in the OSR dataset (selected for their predictivity toward COVID-19). The blue and orange areas correspond to negative and positive cases, respectively.

The missing data rate for each of the examined features is reported in Table 1. In order to reduce the bias due to imputation, we discarded all the features with a missing data rate greater than 75%. Thus, among the 1,624 cases in the OSR dataset, 1,189 (73%) cases had at least 75% of the attributes; while 1,324 (82%) cases had complete data for the CBC features.

From the complete *OSR dataset*, we obtained two other datasets by selecting two relevant subsets of the features (thus, the three datasets share the same set of patients):

1. A dataset consisting of the 34 features under the column header “COVID-specific features” (see Table 1), denoted as the *COVID-specific dataset*.
2. A dataset consisting of the 21 features under the column header “CBC features” (see Table 1), denoted as the *CBC dataset*.

#### External datasets

In addition to the previously described datasets, all obtained from the OSR dataset, we considered two external datasets for the internal–external validation and for the external validation of the models.

The first dataset, the Istituto Ortopedico Galeazzi (*IOG) dataset*, was obtained from blood samples collected at the ED of the IOG of Milan between March 5, 2020, and May 26, 2020, and encompassed the parameters under the “COVID-specific features” column header (see Table 1). Notably, this hospital specializes in the diagnosis and treatment of musculoskeletal disorders and was not considered a destination of choice during the acute phase of the pandemic in the Milan area. Therefore, the patients were presumably of a different severity and were admitted for other reasons than pulmonary conditions with respect to OSR. The IOG dataset consisted of a total of 58 cases, 29 with negative swab results and 29 with positive swab results, and with the same features as the *COVID-specific dataset*. For the IOG and OSR, different instruments were used for the CBC and biochemical parameters; in particular, the iSysmex XN-2000 system was used instead of the Sysmex XE 2100 system for CBC counts and the Atellica® CH Analyzer (Siemens Healthineers) was used instead of the Roche COBAS 6000 system for the biochemical parameters.

The second dataset (the *2018 dataset*) was obtained from blood samples collected at the OSR in November 2018 from 54 randomly chosen patients. These were obviously negative for COVID-19: 20 (37%) of them were specifically chosen to act as confounding cases, as they exhibited pneumonia-like symptoms.

### Machine Learning Experimental Design

We implemented a four-step pipeline for ML model development encompassing imputation, data normalization, feature selection, and classification. The data analysis pipeline was implemented in Python (version 3.7), using the numpy (version 1.19), pandas (version 1.1) and scikit-learn (version 0.23) libraries. For imputation, the multivariate k-nearest neighbors algorithm was used (26), with k = 5. For feature-selection, the recursive feature-elimination algorithm was used (27). The optimal features to select were determined through hyper-parameter optimization. For classification we evaluated five different algorithms: Random Forest (RF), naive Bayes (NB), logistic regression (LR), support vector machine (SVM), and k-nearest neighbors (KNN). We specifically evaluated these algorithms as all have been shown to achieve state-of-the-art performance on tabular data (28) and, at least to some degree (for example using feature-attribution methods), interpretable (29). The hyper-parameters of the different classification algorithms are reported in Suppl. Table 1. All hyper-parameters were optimized automatically using a grid search approach.

In regard to model selection, training and evaluation, we performed a two-step procedure to minimize the risk of over-fitting: first, the dataset was split into a training set (80% of the instances) and a hold-out test set (20% of the instances), using a stratified procedure; second, hyper-parameter optimization was performed (on the training set) through 5-fold stratified cross-validation grid search and using AUC as reference measure; third, the models were trained and calibrated on the whole training set; finally, the calibrated models were evaluated on the hold-out test set in terms of accuracy, sensitivity, specificity, AUC, and the Brier score (30) (a standard metric to measure the models’ calibration, with a lower score being better). In all stages of model development, the randomization was controlled in order to ensure repeatability of the experiments.

For each model class, we considered two versions: a standard one, and the three-way version (a model that abstains from prediction when the confidence score is below 75%) (31). For each of these two versions, the model selection, training and evaluation pipeline was implemented for each of the three datasets mentioned above (the *OSR dataset, COVID-specific dataset*, and *CBC dataset*).

The *IOG dataset* and the *2018 dataset* were used, respectively, for the internal–external and external validation of the models developed for the *COVID-specific dataset* and the *CBC dataset*.

The internal–external validation procedure—the purpose of which was to evaluate the models’ ability to generalize to a new setting when provided with a limited quantity of new data—was implemented using a bootstrap-based approach (see Supplementary Materials - Implementation of the Internal-External Validation).

The external validation procedure—the purpose of which was to test both the specificity of the developed models and their ability to identify potential suspect cases—was implemented by training the best models found for the *COVID-specific dataset* (respectively, the *CBC dataset*) on the combined dataset that also consisted of the *IOG dataset* and then evaluating the trained models against the *2018 dataset*.

The combined dataset consisting of the *COVID-specific* (respectively, the *CBC dataset*) and the *IOG* datasets were also used to evaluate the sensitivity and specificity for symptomatic and asymptomatic patients separately: in this case, the models were retrained after deletion of the Suspect feature (to avoid bias) and the re-trained models were then evaluated on symptomatic and asymptomatic patients (both from the test set) separately.

## Results

The results of the ML models on the three datasets (OSR, COVID-specific, CBC) are reported in Table 2.

**Table 2.** Results for the models trained using the *OSR dataset*, the *COVID-specific dataset and* the *complete blood count (CBC) dataset*. The first value refers to the standard version, the second value to the three-way version. The last column reports on the coverage of this latter model; that is, the proportion of data for which the classifier makes a prediction with at least a 75% confidence score.

| Dataset | Model | Accuracy | Sensitivity | Specificity | AUC | Brier<sup>1</sup> | Coverage for the 3-way version (75% confidence) |
| --- | --- | --- | --- | --- | --- | --- | --- |
| OSR dataset | Logistic Regression | 0.86/0.92 | 0.88/0.95 | 0.84/0.90 | 0.86/0.95 | 0.13 | 0.76 |
|  | Naive Bayes | 0.85/0.87 | 0.82/0.83 | 0.88/0.90 | 0.85/0.91 | 0.12 | 0.94 |
|  | KNN | 0.83/0.89 | 0.76/0.82 | 0.90/0.95 | 0.83/0.90 | 0.12 | 0.72 |
|  | Random Forest | **0.88/0.93<sup>2</sup>** | 0.86/0.92 | **0.91/0.94** | **0.90/0.94** | **0.10** | **0.70** |
|  | SVM | **0.88/0.91** | **0.89/0.92** | 0.87/0.90 | 0.88/0.94 | 0.11 | 0.77 |
| COVID – specific dataset | Logistic Regression | 0.83/0.87 | **0.85/0.89** | 0.82/0.85 | 0.83/0.88 | 0.14 | 0.70 |
|  | Naive Bayes | 0.83/0.88 | 0.84/0.85 | 0.83/0.91 | 0.83/0.91 | 0.13 | 0.76 |
|  | KNN | **0.86/0.90** | 0.80/0.85 | **0.92/0.94** | **0.87/0.94** | **0.11** | **0.81** |
|  | Random Forest | 0.84/0.89 | 0.84/0.92 | 0.84/0.87 | 0.84/0.92 | 0.12 | 0.82 |
|  | SVM | **0.86/0.87** | 0.83/0.83 | 0.89/0.91 | 0.86/0.93 | 0.12 | 0.74 |
| CBC dataset | Logistic Regression | 0.74/0.80 | 0.70/0.78 | 0.79/0.83 | 0.74/0.85 | 0.18 | 0.60 |
|  | Naive Bayes | 0.78/0.83 | 0.74/0.79 | 0.82/0.87 | 0.78/0.88 | 0.16 | 0.69 |
|  | KNN | **0.86/0.90** | 0.82/0.84 | **0.89/0.95** | **0.86/0.89** | **0.13** | 0.76 |
|  | Random Forest | 0.83/0.90 | **0.84/0.92** | 0.82/0.87 | **0.86/0.91** | **0.13** | 0.68 |
|  | SVM | 0.77/0.91 | 0.70/0.90 | 0.82/0.92 | 0.76/0.92 | 0.14 | 0.70 |

<sup>1</sup> Brier score, the lower it is, the better it is.
<sup>2</sup> The best value, for each score, is denoted in bold

The receiver operating characteristic (ROC) curves of the best model (in terms of the highest AUC) for each of the three datasets is reported in Figure 3. The ROC curves for all models (on each of the three datasets) are reported in Suppl. Figures 1, 2 and 3. The feature importance scores—which were computed in order to enable the interpretability of the developed models— are reported in Figure 4 and in Suppl. Figure 13 for the best model of each of the three datasets. The positive predictive value (PPV)-sensitivity curves are reported in Suppl. Figures 4, 5 and 6, while the calibration curves are reported in Suppl. Figures 7, 8, and 9, and the PPV/NPV prevalence curves are reported in Suppl. Figures 10, 11 and 12.

**Figure 3.**
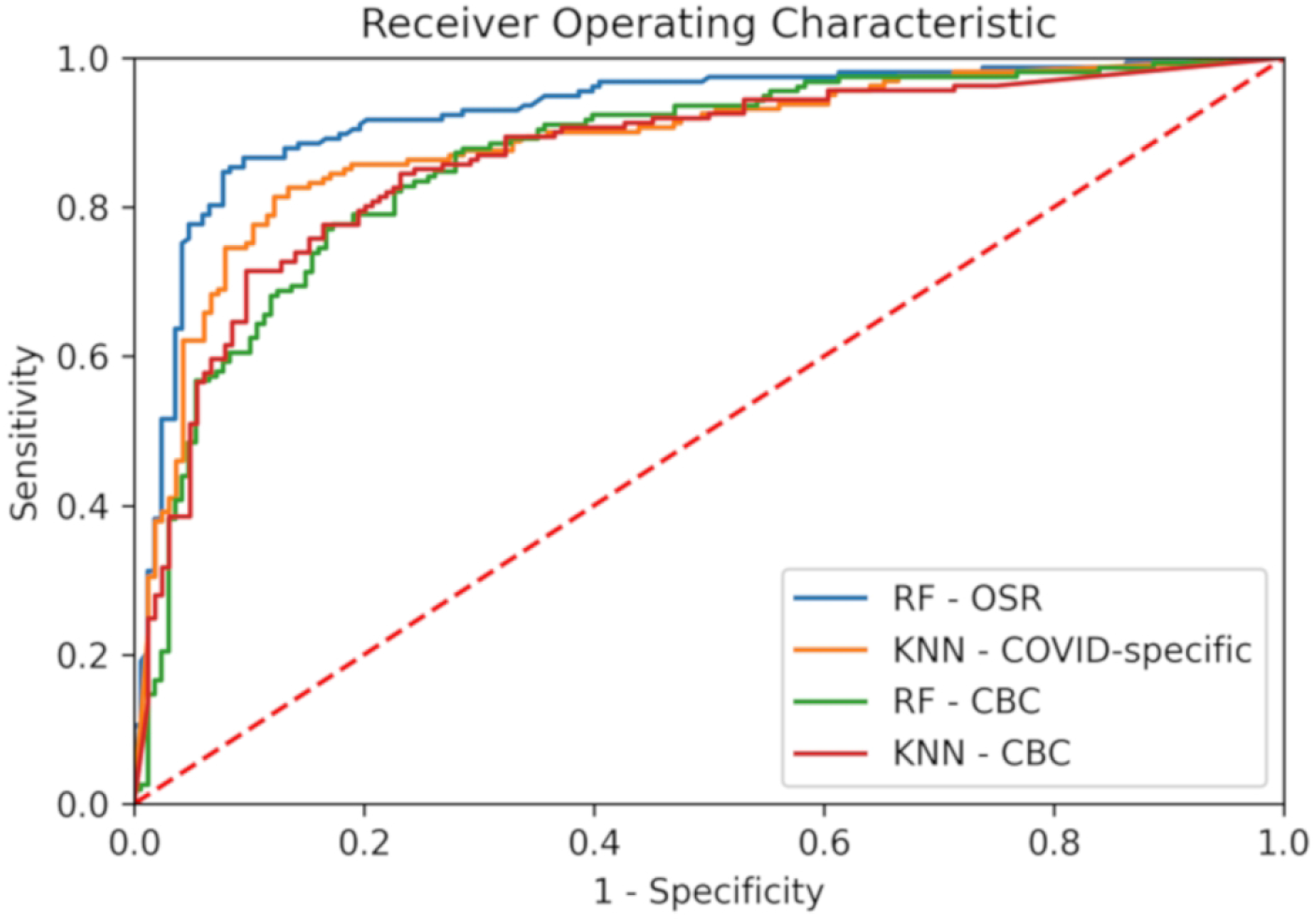
Receiver operating characteristic curves for the best models (in terms of AUC), for each of the three considered datasets (OSR, COVID-specific, CBC). For the CBC dataset we report the ROC curve for both RF and KNN as they had equal AUC (see Table 2).

**Figure 4.**
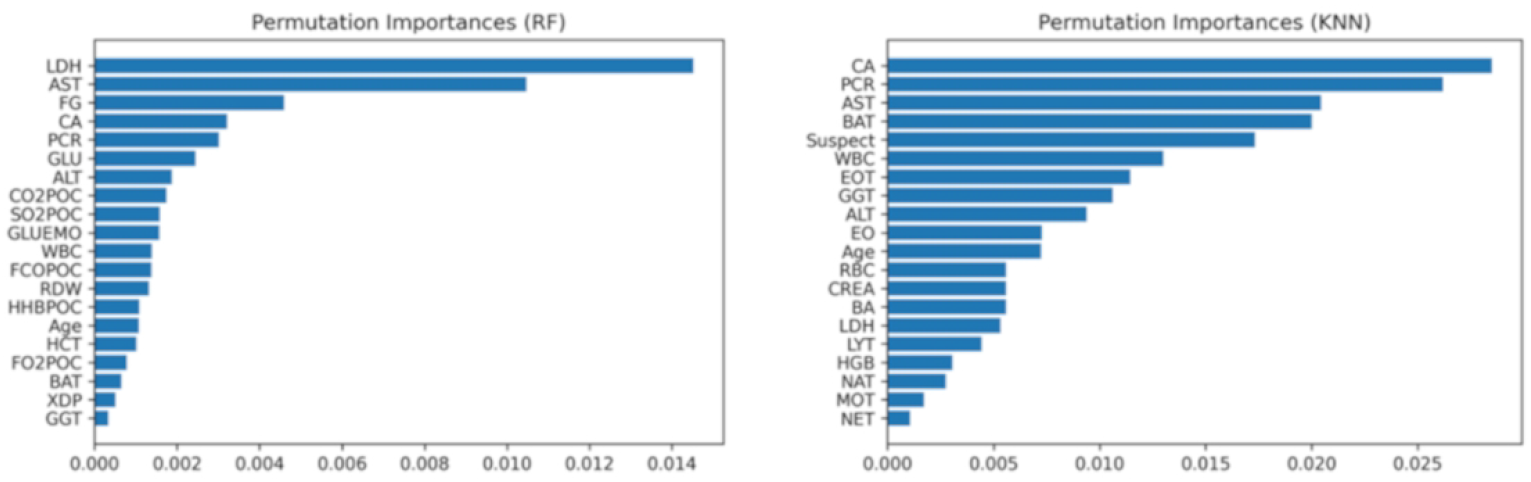
Feature importance scores for the random forest algorithm trained using the OSR dataset (on the left) and the k-nearest neighbors’ algorithm trained using the COVID-specific dataset (on the right).

The results for the internal–external validation and the external validation (specificity only) procedures are reported in Table 3. In this table we highlight the results of the models that obtained the best performance in the internal validation (KNN for the COVID-specific dataset; and both KNN and RF for the CBC dataset). Specifically, in the first 4 columns we report the results of the internal-external validation (in terms of accuracy, sensitivity, specificity and AUC), while in the last column we report the results of the external validation (in terms of specificity). In Table 3, we also report on the performance of the models for asymptomatic patients and symptomatic patients, as described in the Methods section.

**Table 3.**
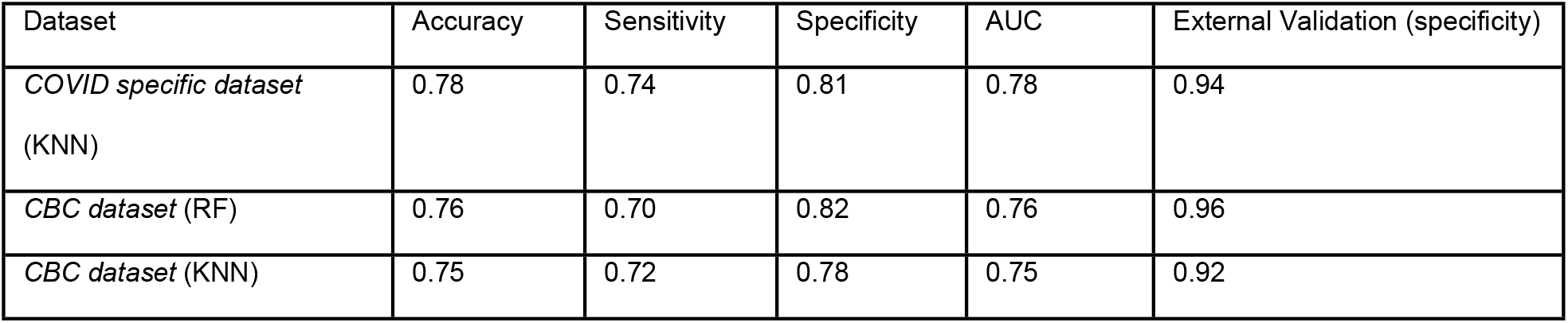
Results for the best models for the internal–external and external validation procedures. For each of the two features sets considered for the internal-external and external validation (namely, COVID-specific and CBC) we report the performances of the best models on the internal validation: namely, KNN for COVID-specific; and both Random Forest and KNN for CBC. The first four columns report on the results on the internal-external validation, while the last column reports on the results of the external validation.

## Discussion

The unprecedented worldwide public health emergency caused by the COVID-19 pandemic has motivated different research groups to develop ML applications with the aim of automating—at least partially—the diagnosis or screening of COVID-19.

Nonetheless, only a few studies have focused on the development of ML models based on routine blood exams. Formica et al.(13) developed a CBC-based ML model, reporting 83% sensitivity and 82% specificity; however, the analysis was based on a small sample (171 patients) collected in a limited time frame (between March 7 and March 19, 2020). Banerjee et al. (32) applied ML methods to a public dataset of CBC data encompassing 598 cases of which only 39 cases were COVID-19 positive; the authors report good specificity (91%) but very low sensitivity (43%), thus making the proposed model unsuitable for early detection tasks. Further, this work presents some major limitations affecting replicability and generalizability, as the authors do not provide any information regarding how the values of the considered features were measured (analytical instruments, analytical principle, and units of measurement). Avila et al. (33) used the same dataset considered in (32) to develop a Bayesian model, reporting 76.7% sensitivity and specificity. Notably, the authors report a number of complete instances (510) which is different from that reported in (32). Joshi et al. (34) developed a logistic regression model trained using CBC data on a dataset of 380 cases, reporting good sensitivity (93%) but low specificity (43%).

More in general, a recent critical survey (5) raised some concerns about these and other evaluated studies (most of which have not yet undergone peer-review), noting the possibility of high rates of bias and over-fitting, and little compliance with reporting and replication guidelines (18).

Finally, a recent study Yang et al. (35), considered the development of a Gradient Boosting model on a set of 3,356 patients (42% COVID-19 positive) using a set of 27 parameters encompassing both blood count and biochemical parameters, achieving 0.85 AUC, and also reporting a comparable result (AUC 0.84) for validation on an external dataset. This work can be viewed as similar but complementary with respect to the results that we report, both in terms of considered features and used laboratory instrumentation (the authors used the UniCel DXH 800 analyzer for the CBC features, and Siemens ADVIA XPT analyzers for biochemical parameters). Indeed, compared with the parameters considered in this study, the authors of (35) considered albumin, total protein, magnesium, ferritin and globulin; but lacked a set of parameters (some of which known to be significantly altered in COVID-19 patients), such as creatinine (CREA), aspartate aminotransferase (AST), alanine aminotransferase (ALT), Gamma-glutamyl transferase (GGT), Creatin-kinase (CK), Potassium (K), Interleukin 6 (IL6), NT-proB-type Natriuretic Peptide (ProBNP), total (BilT) and direct (BilD) Bilirubin, all coagulation tests, hemogasanalysis and CO-Oxymetry parameters. We think that this complementarity in the two studies could lend support to the usefulness of blood tests as an alternative approach for COVID-19 diagnosis.

To overcome the limitations of the above models, we applied the ML methodology to routine blood examination outcomes, which are usually available for inpatients and for patients admitted to the ED in shorter time frames and at much lower cost than both molecular tests and radiological exams. In this endeavor, we addressed three subtasks:1) detecting COVID-19 from a full battery of hematochemical tests, commonly collected from suspected respiratory tract disease patients (OSR dataset); 2) detecting COVID-19 from only a restricted subset of parameters known to be altered in COVID-19 patients (COVID-specific dataset); and 3) detecting COVID-19 from a very small subset of hematological parameters (CBC and the WBC differential) representing the basic routine blood examinations, usually also available in low-resource settings (CBC dataset). For each of the datasets described above, we applied five different models that were selected from among those that are more frequently adopted in medical ML.

These models achieve COVID-19 detection in different ways and exhibit good performance, although they are associated with different sensitivities and specificities. This makes them good candidates for embedding in an online service (which we are currently developing) in which doctors can specify their preferences (with respect to greater sensitivity, greater specificity, or a balanced performance (36) and needs according to their diagnostic purpose (i.e., screening, triage, or a secondary diagnosis), and thus gain an indication from the optimal model. In addition, the users can decide whether they want an indication from the system—irrespective of the confidence in the advice given—or if they would prefer only to be advised about high-confidence indications, as the three-way approach allows for. This approach was specifically developed to mitigate the risk of automation bias and the odds of machine-induced errors (31).

With respect to the patterns used to discriminate between positive and negative cases, the ML models identified, as the most predictive features, those parameters that are known to be significantly altered in COVID-19 patients (5,37-38) (see Figure 4 and Suppl. Figure 13). For instance, when applied to the OSR and COVID-specific datasets, the models identify lactate dehydrogenase (LDH), AST, C-reactive protein (CRP), and calcium as the most important features, while all the models also reported WBC and its corresponding differential as important. Also the patients’ age was reported by the models to be a significant predictor, which is consistent with the literature (5), where it was also found to be a significant predictor not only for prognostic, but also for diagnostic tasks. Notably, fibrinogen and cross-linked fibrin degradation products (XDPs), known to be associated with COVID-19 severity (39), were also considered by the model as being among the most important features when applied to the OSR dataset.

With respect to the calibration of the developed models, the good internal calibration of the models can be confirmed by the calibration curves in Suppl. Figures 7–9.

As can be seen in Table 3, the internal–external validation and the external validation procedures also achieved good results. In this respect, it is important to note that the validation procedures involved blood tests performed on different types of analytical instrumentation for the clinical chemistry tests (Siemens instead of Roche), although the CBC standardization was less problematic than the other tests. For this reason, as ML models exhibit poor performance when considering out-of-distribution samples (40), the goal of the internal–external validation process was to assess the capability of the models to generalize across different settings. All of the models showed good performance and, more specifically, good specificity. The models achieved good performances in symptomatic patients (with both the sensitivity and specificity at approximately 80%) and they performed even better in terms of specificity in asymptomatic patients (100% specificity), although the sensitivity was as low as 50% (see Table 3). Nevertheless, considering that the developed ML-based tests were based on low-cost and rapid blood-test examinations, the reported values can be considered good enough, specifically in regard to screening (16).

The external validation procedure also achieved very good results (at around 95% for all models in the standard version), but it should be noted that this only relates to COVID-19-negative patients, and hence, to specificity. Notably, in the external validation process, all five patients for which the models failed had symptoms that were compatible with COVID-19 disease.

As hinted at above, the outputs from our models can be used in different scenarios. They could be used together and combined with the molecular test to obtain a compound test with higher accuracy, and, most importantly, higher sensitivity regarding suspected cases, thus allowing for the identification of a larger number of COVID-19-positive patients so that they can be isolated and treated in a timely manner. Indeed, we can see in Table 3 that the sensitivity in symptomatic patients is adequate for this type of use. In the same vein, the models’ outputs could be used while waiting for the results from other tests, allowing for the timely and prudent management of suspected COVID patients, or in screening and pool-testing scenarios (41) where low accuracy is not a critical problem if a test such as a CBC can be performed frequently (42). In Table 3 we can see that the model that was defined based on the smallest dataset (the CBC dataset) reaches 100% specificity in asymptomatic patients. Consequently, we are planning to use our model for epidemiological purposes on the blood donor population to estimate the prevalence of the condition in the asymptomatic population. On the other hand, the scenarios in which the results from our models replace those of the molecular tests address an emergency need, especially if the time to obtain the molecular test results is too long (due to a high demand for such tests in an outbreak area), if there is a shortage of materials (swabs or reagents) for any supply problem, or in poor health contexts or in contexts where there are serious structural deficiencies (such as in some developing countries or in a geographical area that is, in the meantime, affected by other socio-sanitary and humanitarian emergencies). In these situations where resources are limited and population-wide testing cannot be performed, CBC-based scores may help to pre-evaluate patients and activate COVID-19-specific pathways and molecular testing for patients with high scores independent of symptom severity. In the presence of suspected COVID-19 cases and high scores, logistical management can promptly activate isolation procedures (43).

## Conclusion

All things considered, the ML models that we presented in this article achieved a performance that is comparable, although inferior, to RT-PCR (4), which is the current gold standard for COVID-19 diagnosis. Nevertheless, although our models are less accurate, they aim to be an additional tool available among those that, being much faster and cheaper than the current diagnostic reference tests, can be used for the screening of whole populations. This use can facilitate the shift in testing strategy that, grounding on a faster, although less accurate, identification of infected individuals, is said to have a positive potential in slowing the virus’ spread and contributing for the safe reopening of schools and workplaces (44).

## Data Availability

The datasets collected and used in this study are available from the corresponding author on reasonable request.

## Author Contributions (in alphabetical order)

Giuseppe Banfi: conceived and designed the study, revised, and approved the manuscript

Federico Cabitza conceived and designed the study, analyzed, and interpreted the data, wrote and revised the manuscript, approved the manuscript.

Andrea Campagner: analyzed and interpreted the data, wrote and revised the manuscript, approved the manuscript.

Anna Carobene: conceived and designed the study, provided study materials or patients, collected and organized the data, wrote and revised the manuscript, approved the manuscript

Daniele Ceriotti: provided study materials or patients, revised and approved the manuscript

Alessandra Colombini: provided study materials or patients, revised and approved the manuscript

Elena De Vecchi: provided study materials or patients, revised and approved the manuscript

Chiara Di Resta: collected and organized the data, wrote and revised the manuscript, approved the manuscript

Davide Ferrari: conceived and designed the study, revised and approved the manuscript

Massimo Locatelli: conceived and designed the study, revised and approved the manuscript Eleonora Sabetta: provided study materials or patients, revised and approved the manuscript

## SUPPLEMENTAL MATERIAL

**Suppl. Table 1.** Hyper-parameters of the machine learning models under consideration.

| Model | Random Forest | Naive Bayes | Support Vector Machine | Logistic Regression | k-Nearest Neighbor |
| --- | --- | --- | --- | --- | --- |
| Hyper-parameters | Number of estimators, Maximum tree depth, Split criterion, Maximum number of features | / | Kernel, Maximum polynomial degree, Kernel coefficient, Regularization parameter | Regularization penalty, Regularization parameter | Distance weighting, Nearest Neighbor algorithm, Number of neighbors |

### Supplementary Figures

**Suppl. Figure 1.**
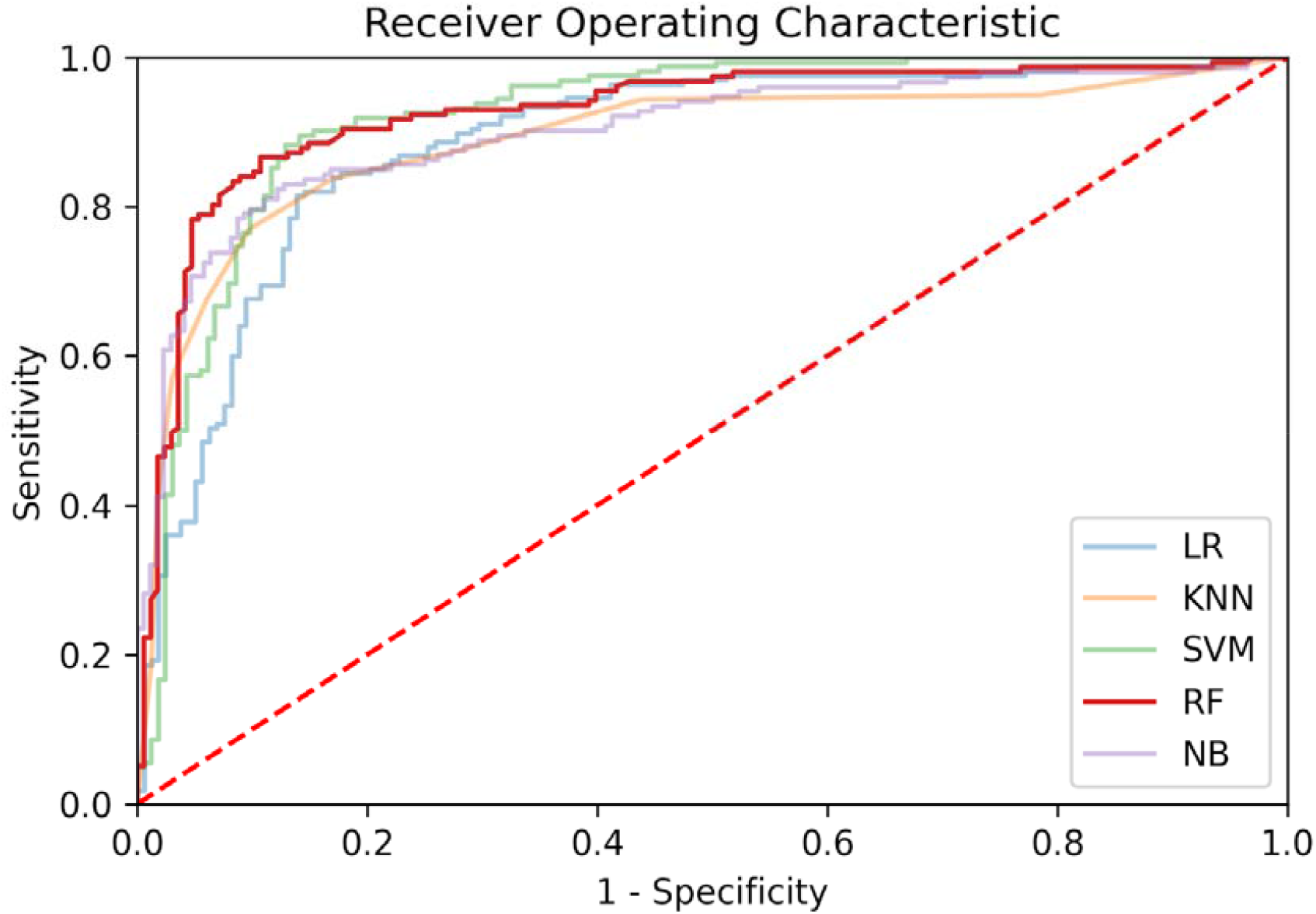
Receiver operating characteristic curves for the models trained using the *OSR dataset*.

**Suppl. Figure 2.**
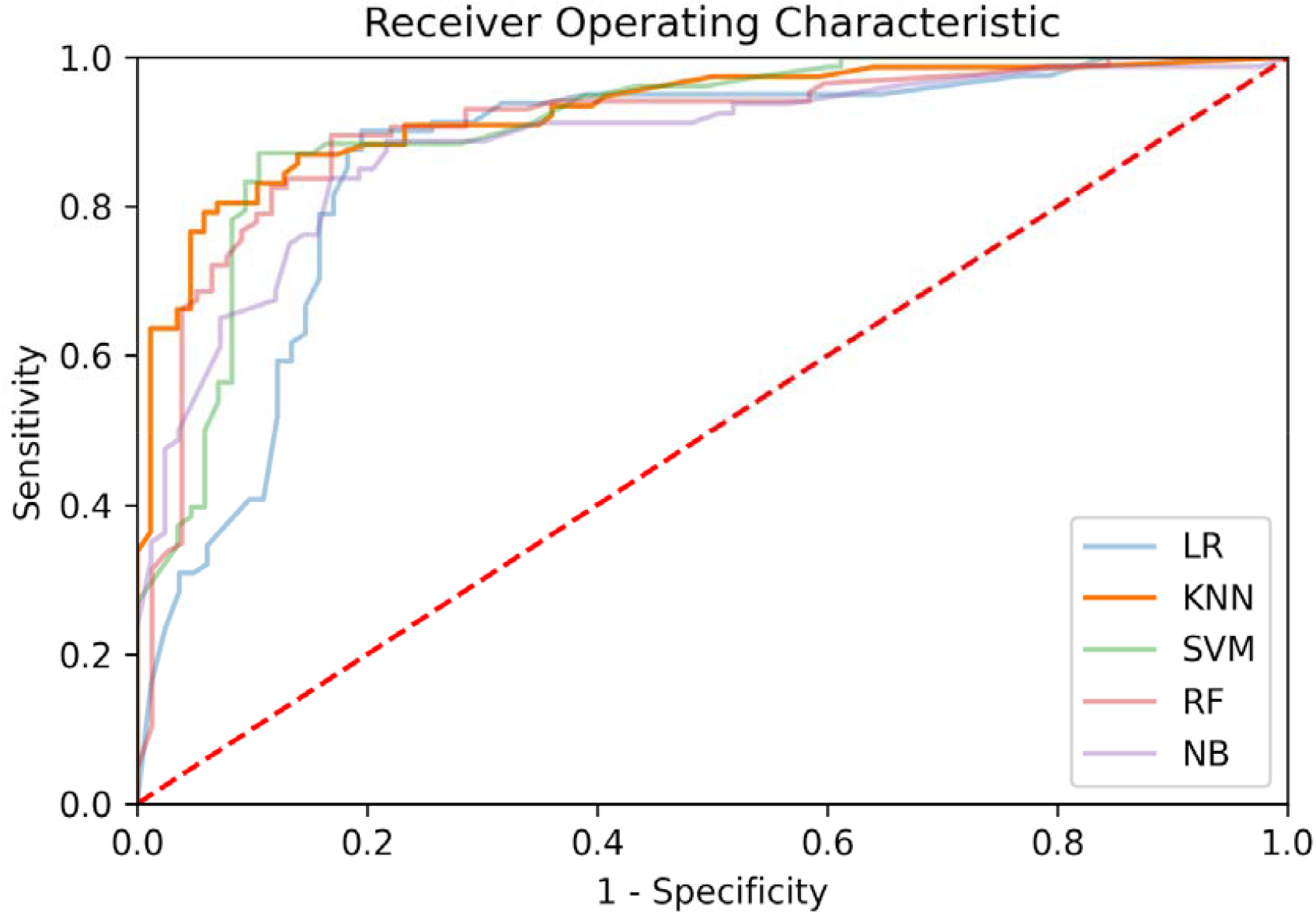
Receiver operating characteristic curves for the models trained using the *COVID-specific dataset*.

**Suppl. Figure 3.**
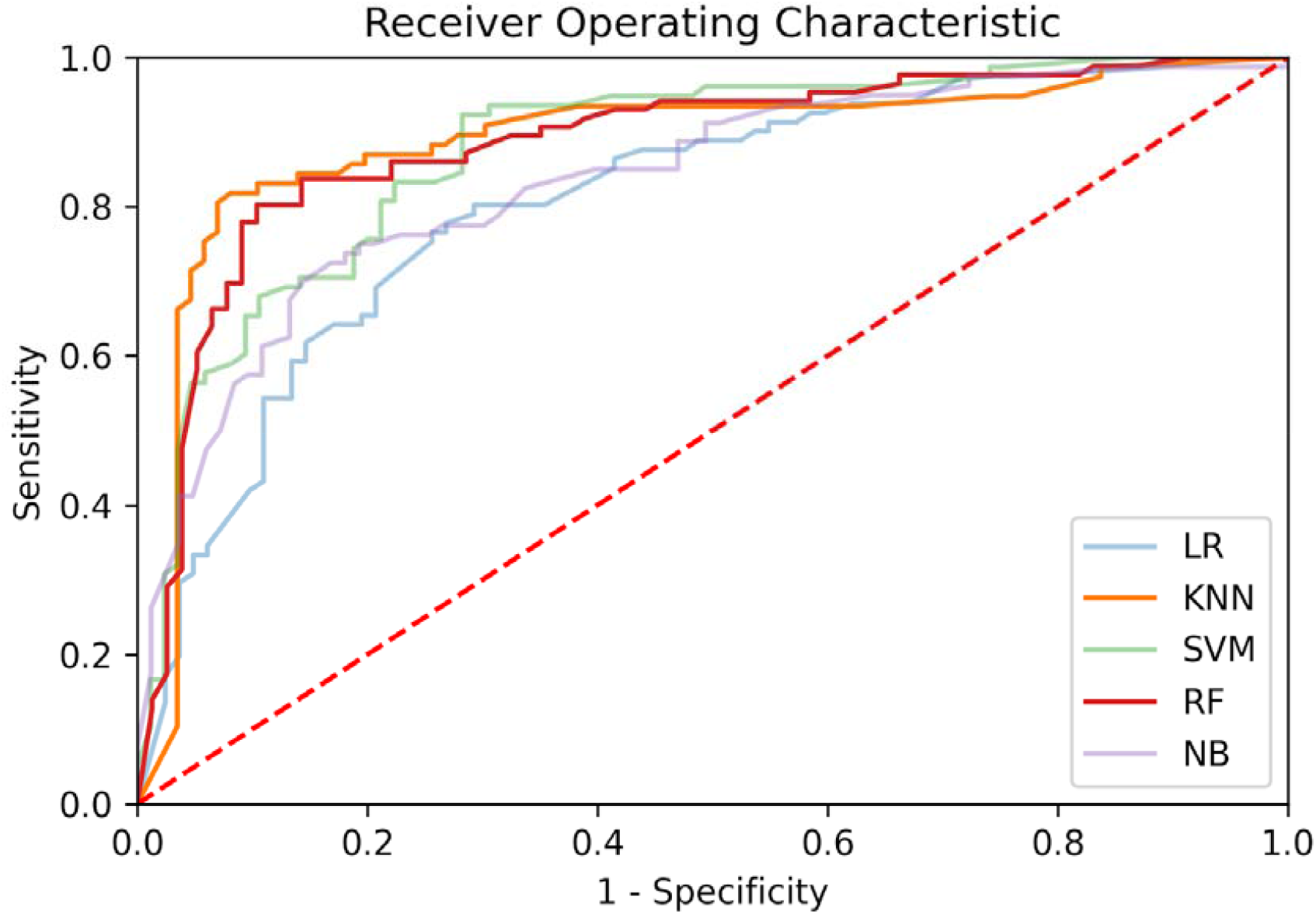
Receiver operating characteristic curves for the models trained using the *CBC dataset*.

**Suppl. Figure 4.**
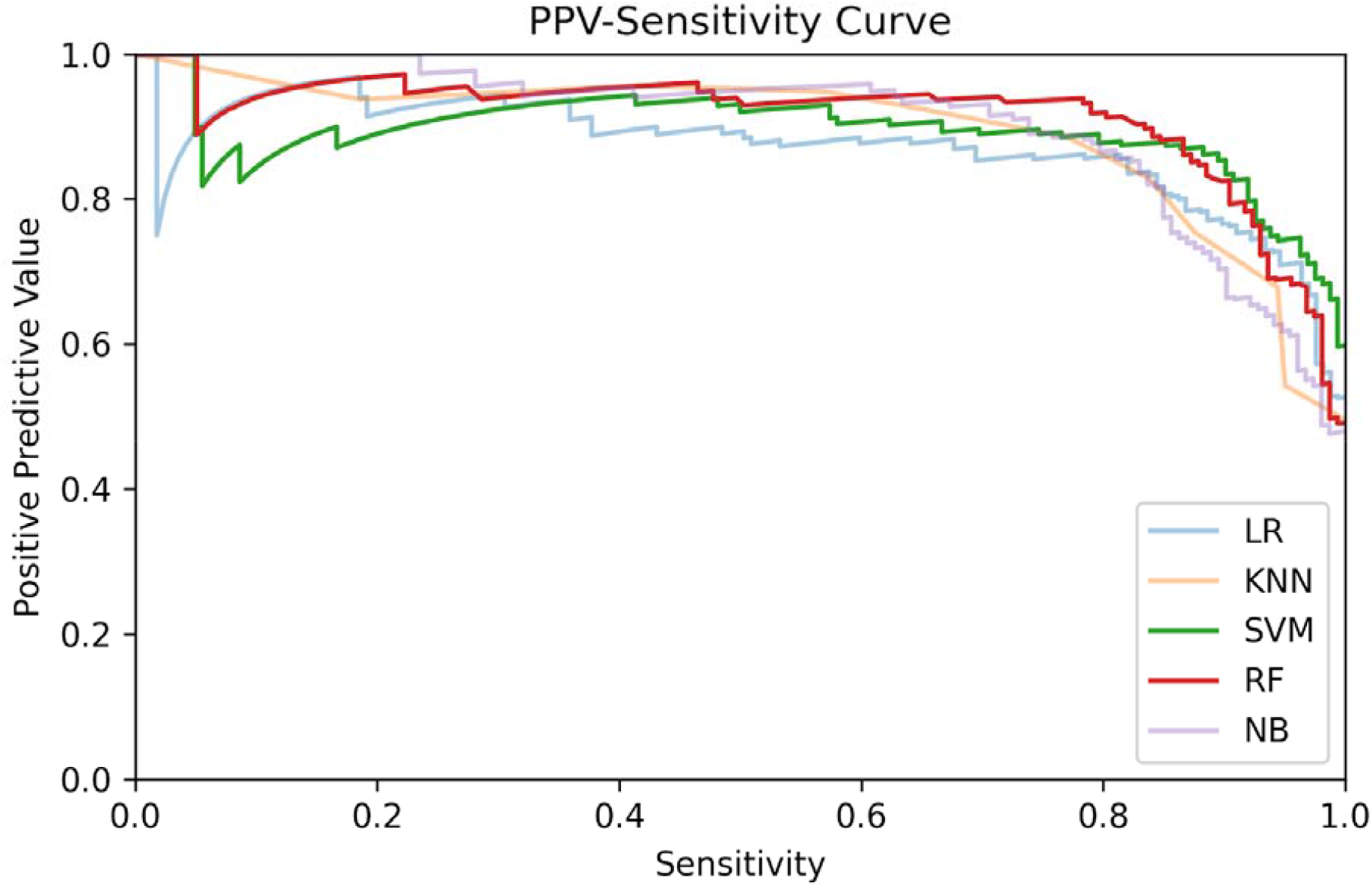
Positive predictive value-sensitivity curves for the models trained using the *OSR dataset.*

**Suppl. Figure 5.**
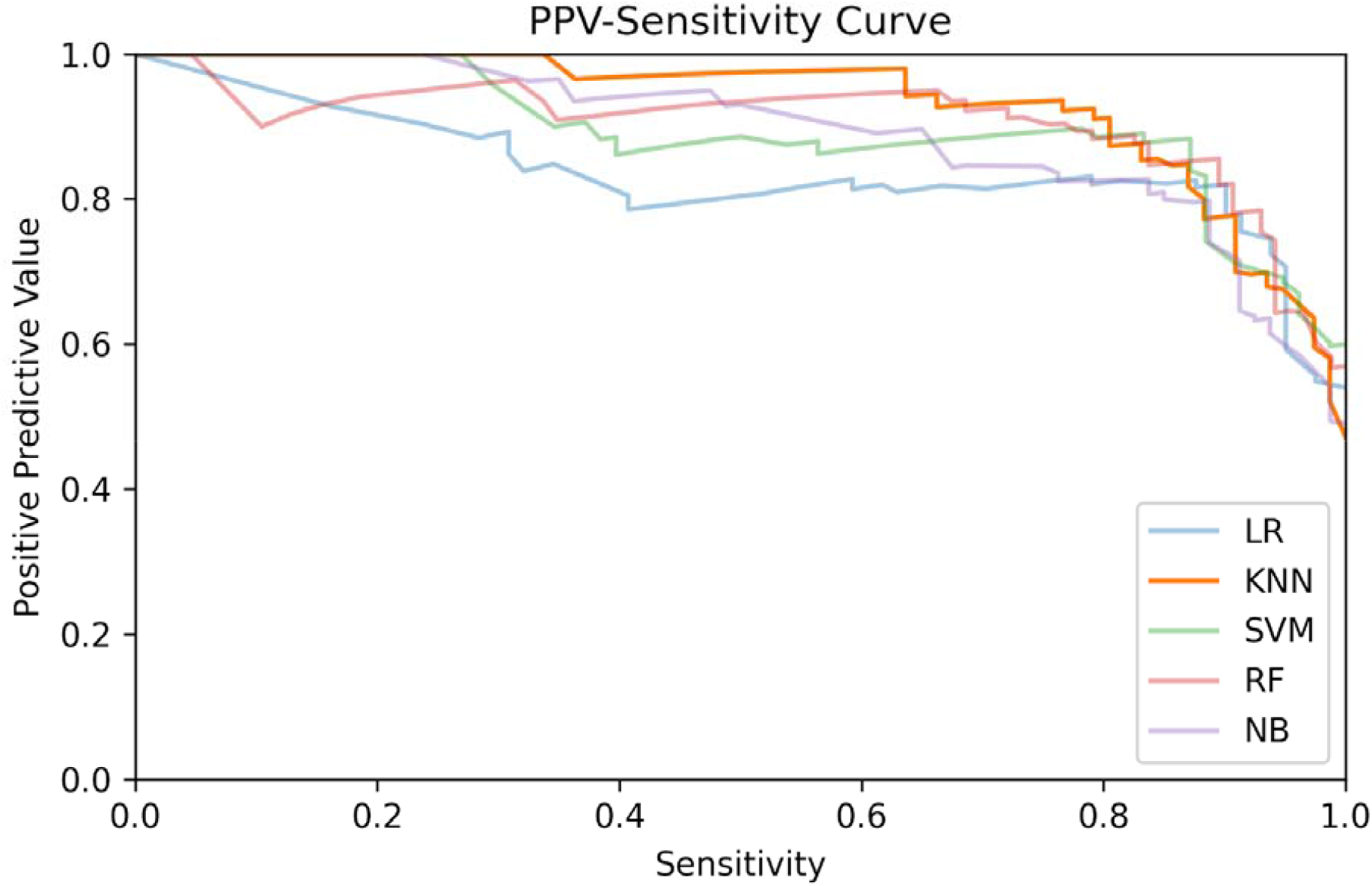
Positive predictive value-sensitivity curves for the models trained using the *COVID-specific dataset*.

**Suppl. Figure 6.**
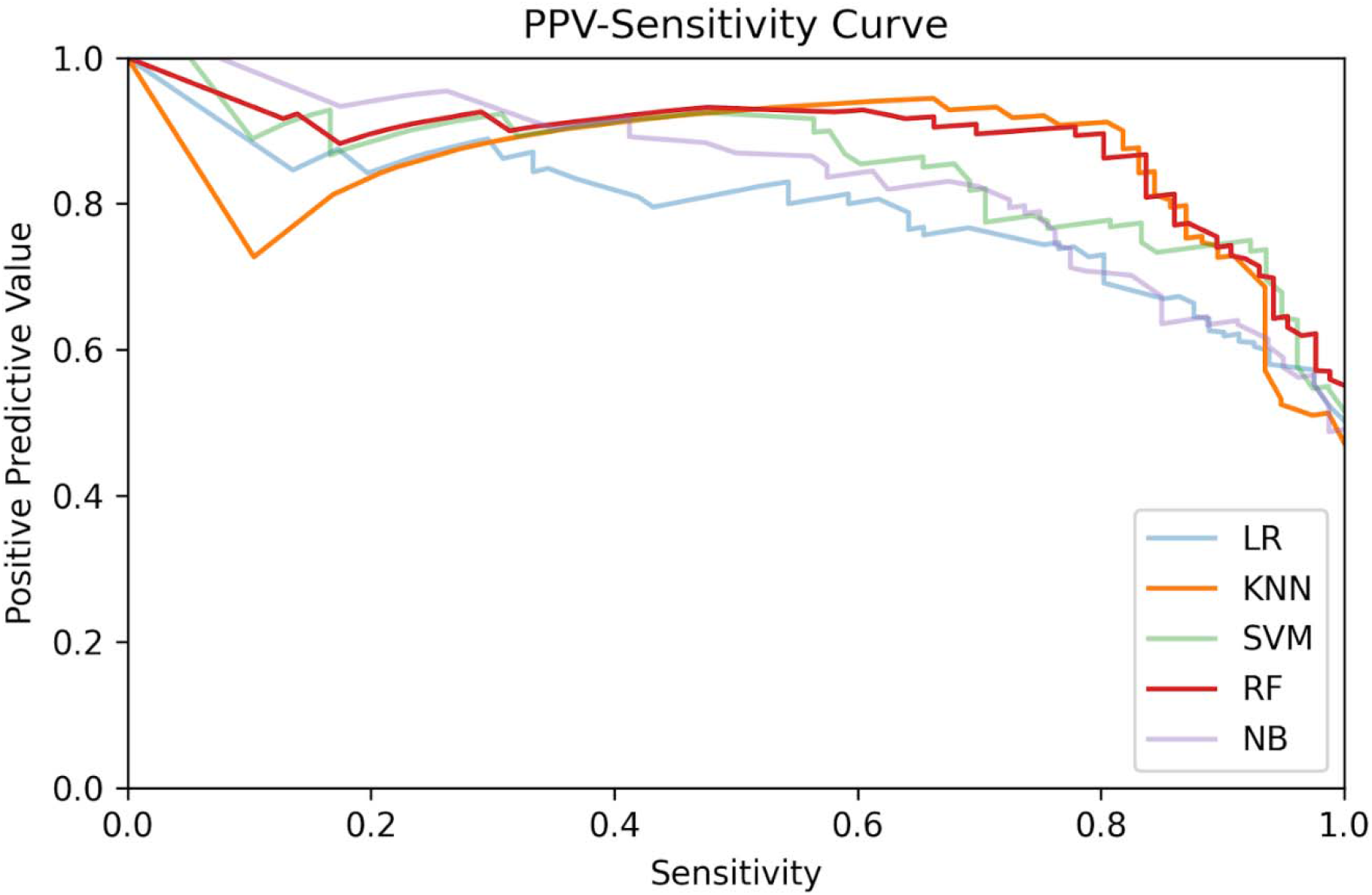
Positive predictive value-sensitivity curves for the models trained using the *CBC dataset*.

**Suppl. Figure 7.**
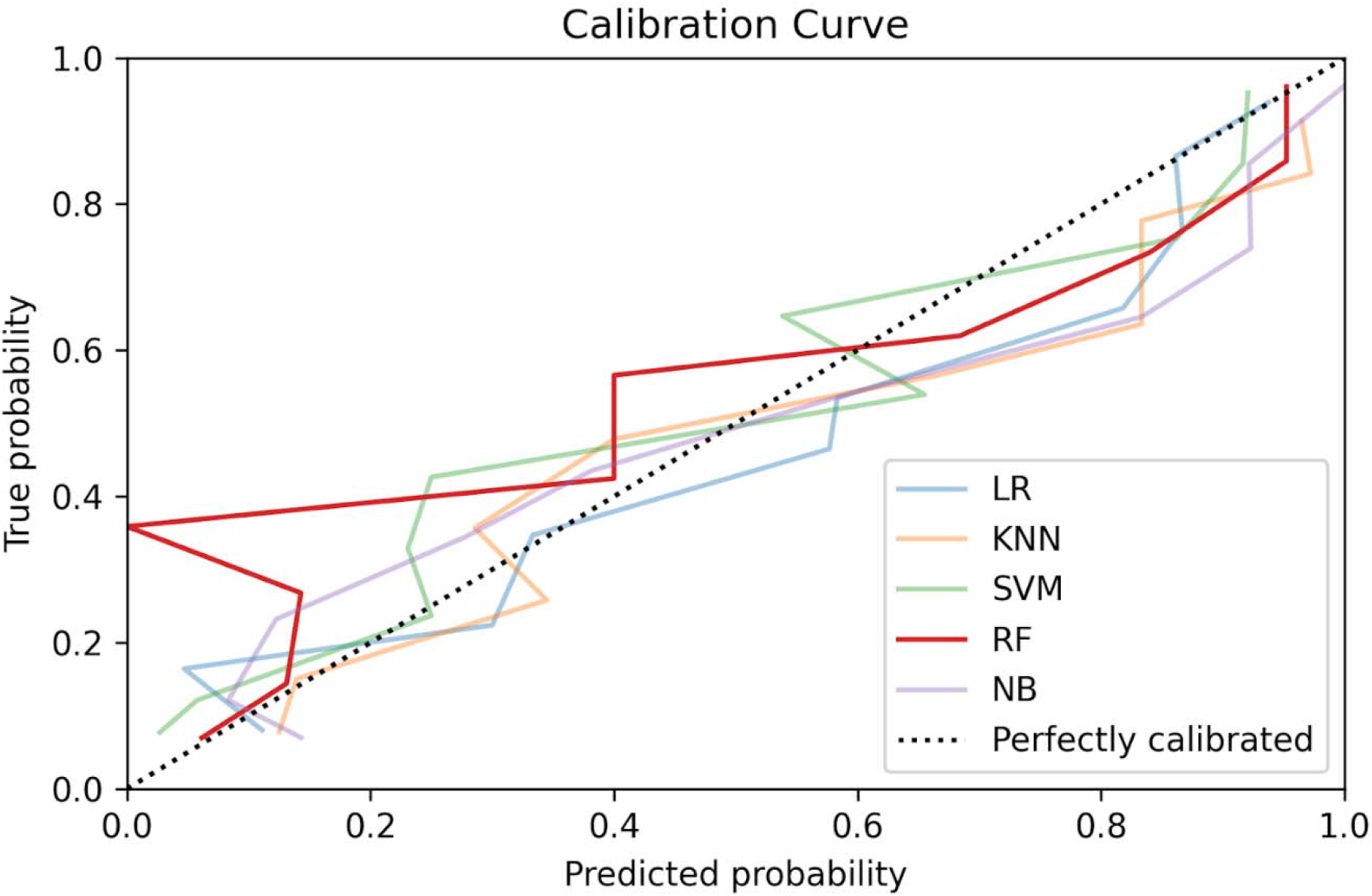
Calibration curves for the models trained using the *OSR dataset*.

**Suppl. Figure 8.**
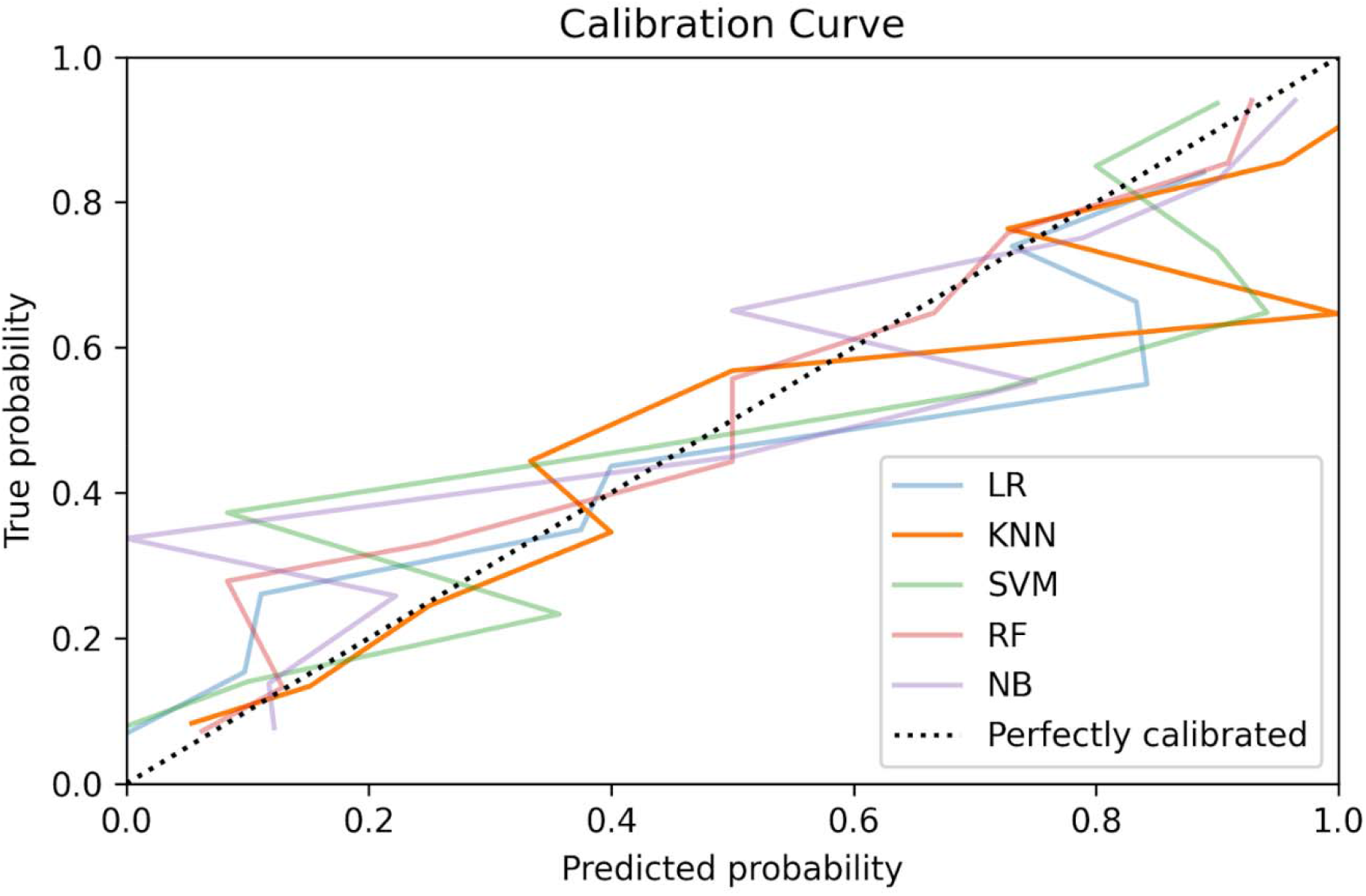
Calibration curves for the models trained using the *COVID-specific dataset*.

**Suppl. Figure 9.**
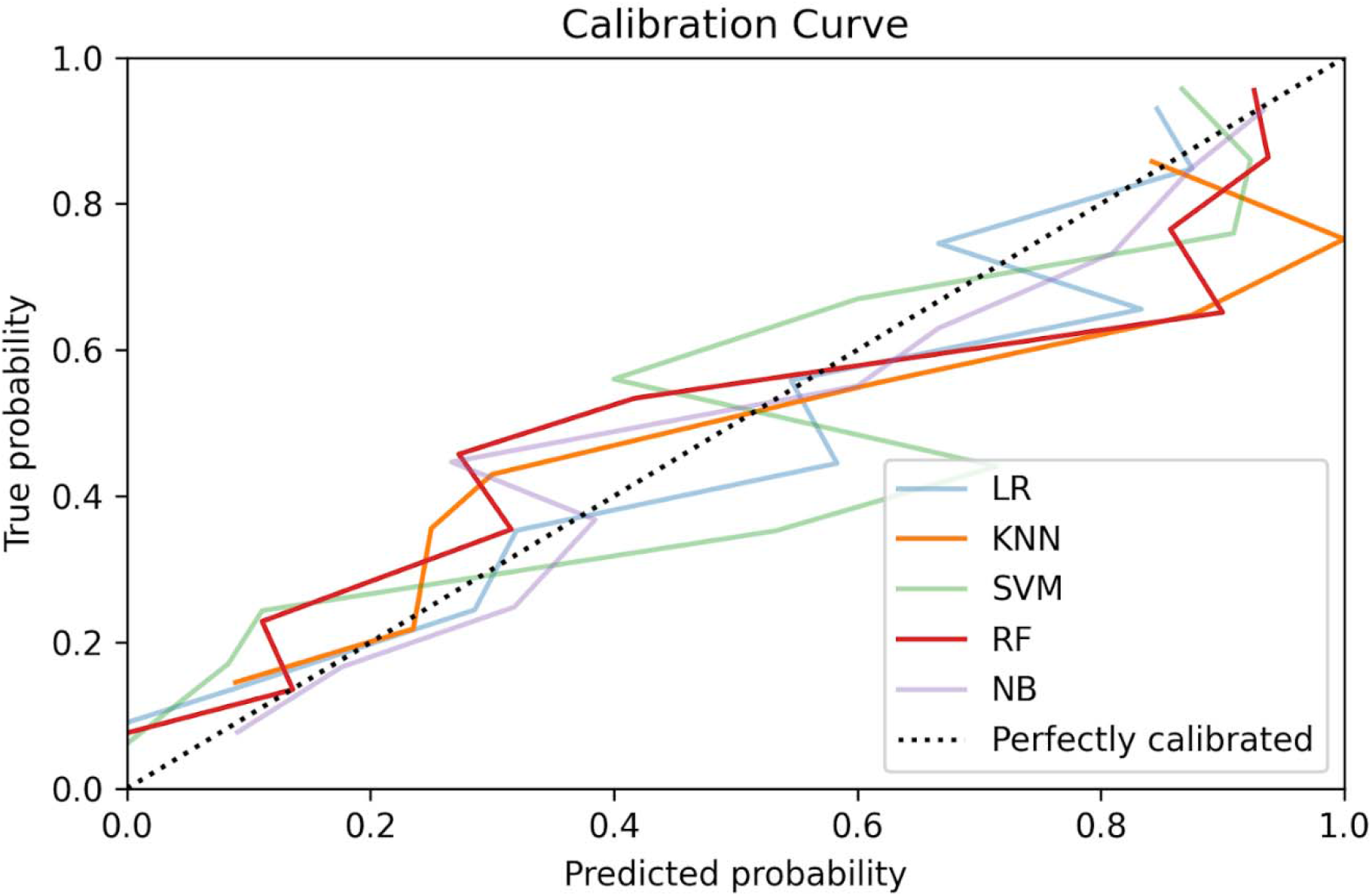
Calibration curves for the models trained using the *CBC dataset*.

**Suppl. Figure 10.**
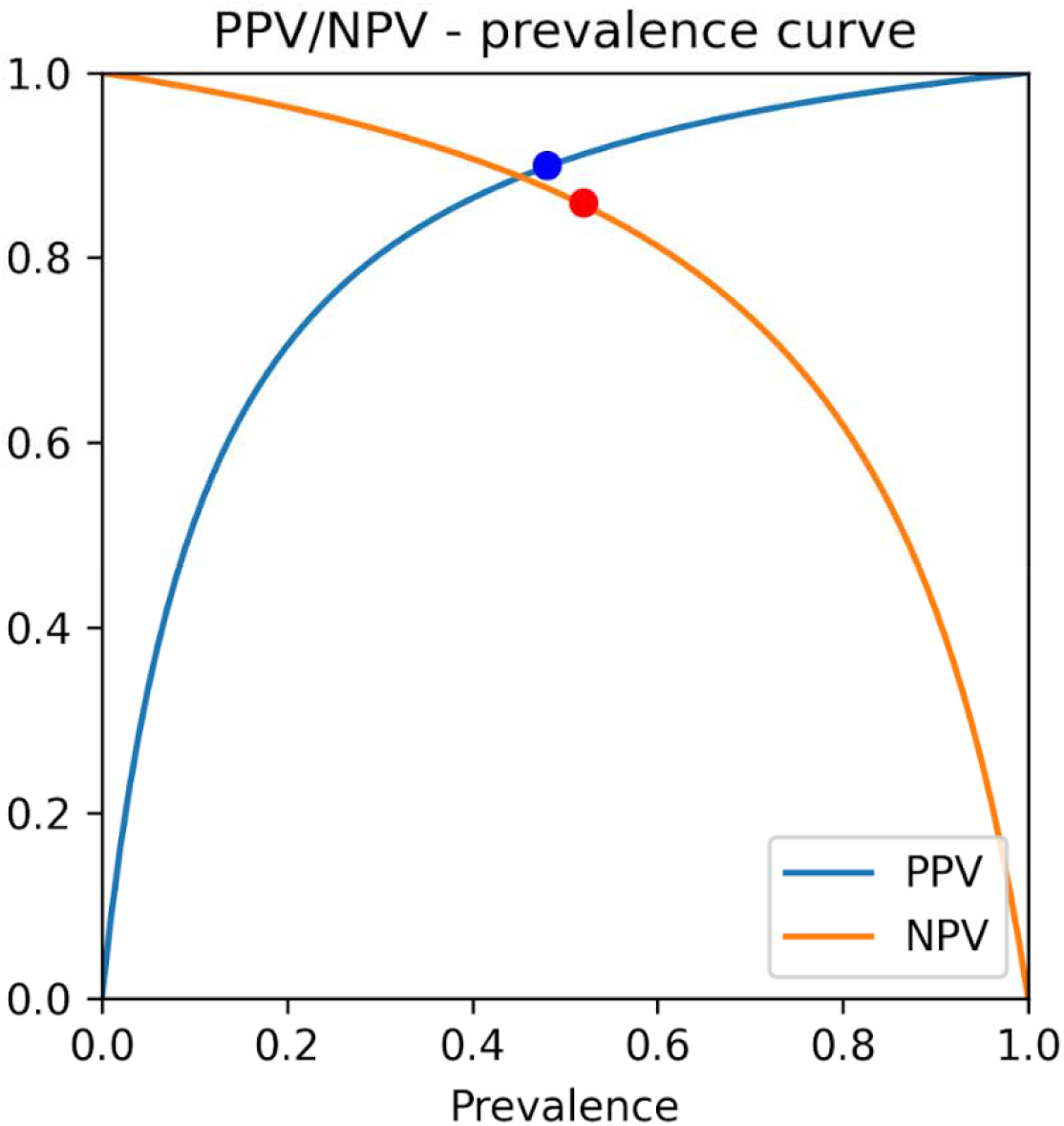
Positive predictive value/negative predictive value-prevalence curve for the random forest algorithm, trained using the *OSR dataset*. The points on the curves indicate the prevalence in the dataset.

**Suppl. Figure 11.**
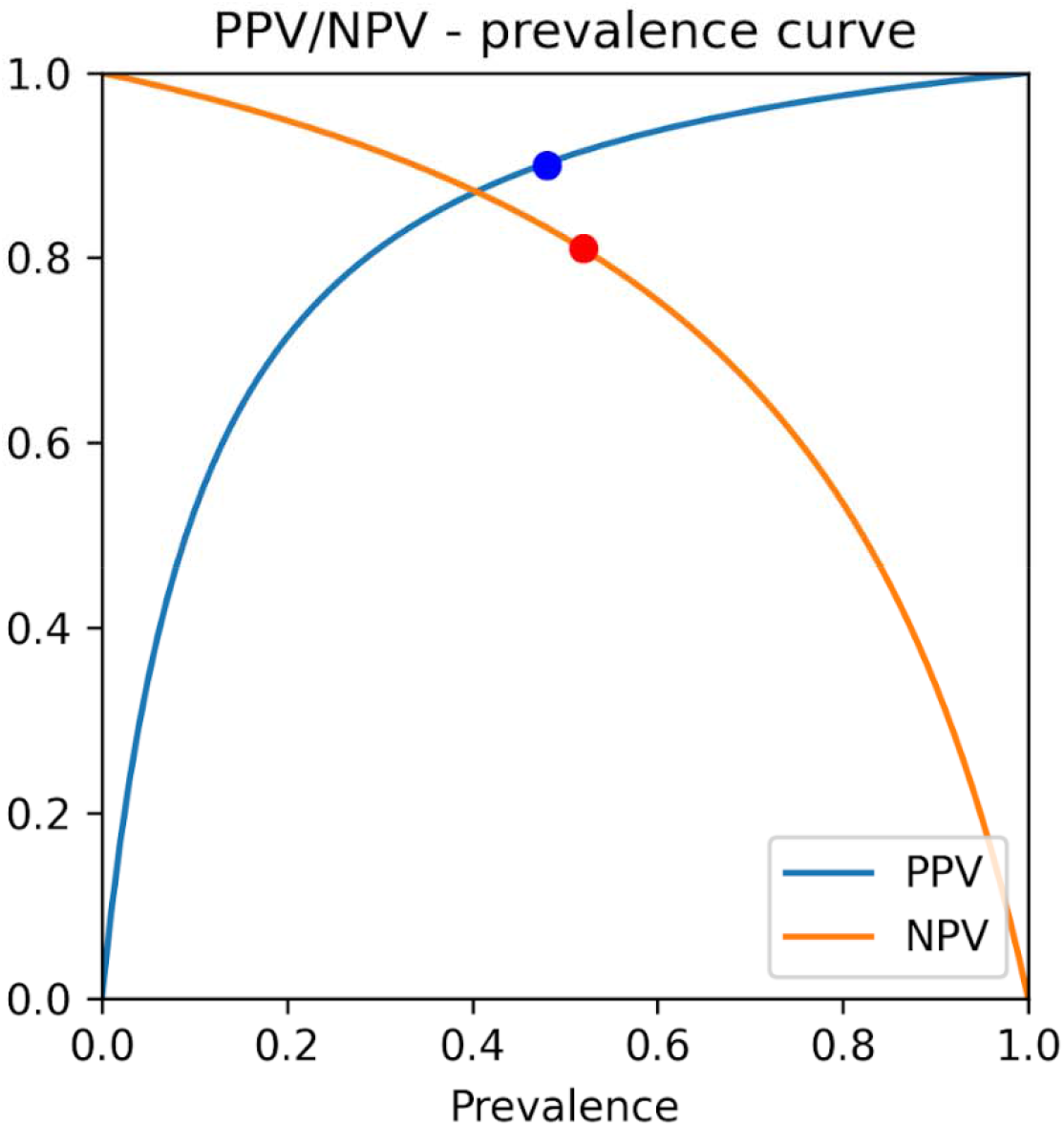
Positive predictive value/negative predictive value-prevalence curve for the k-nearest neighbors’ algorithm, trained using the *COVID-specific dataset*. The points on the curves indicate the prevalence in the dataset.

**Suppl. Figure 12.**
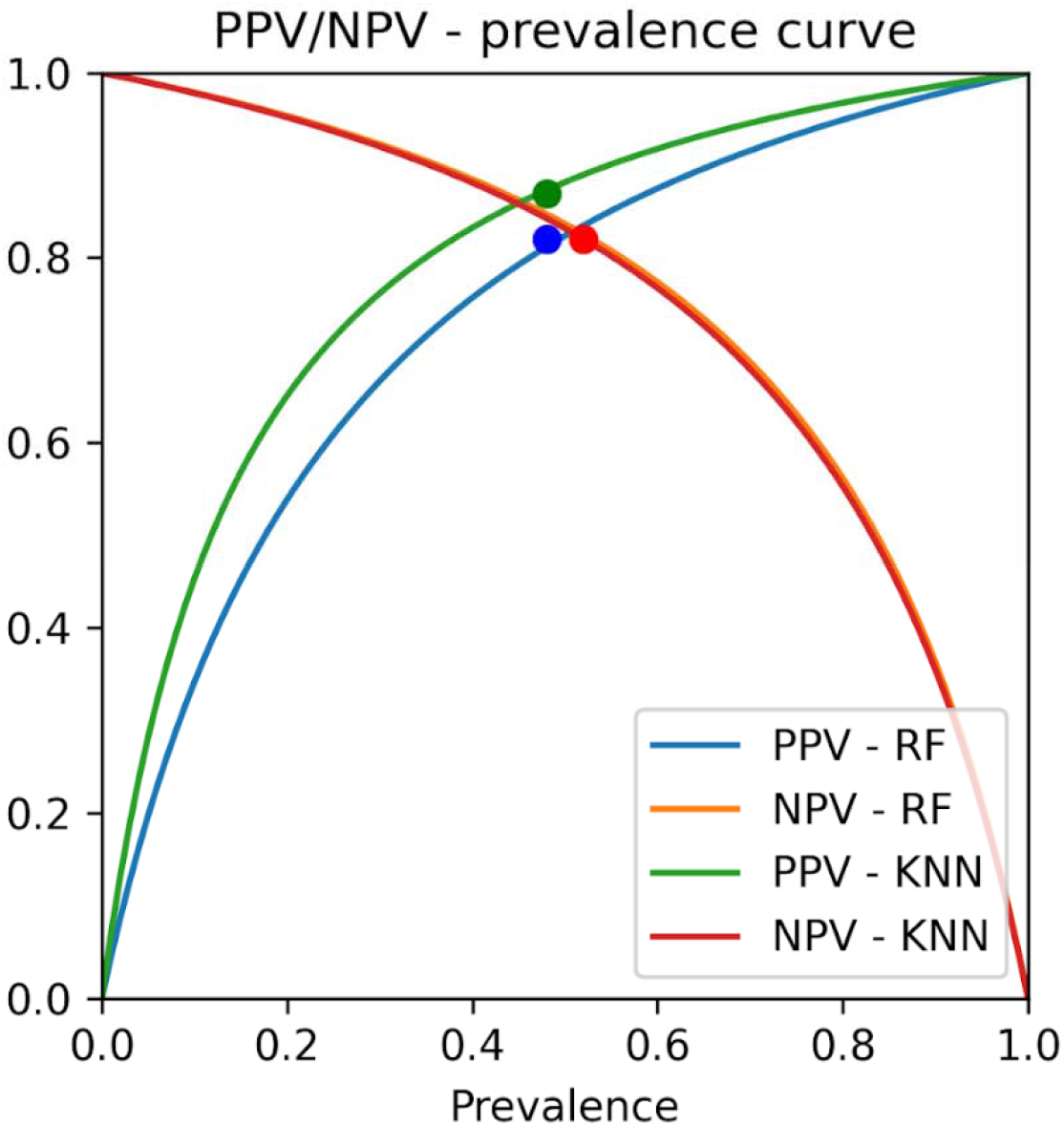
Positive predictive value/negative predictive value-prevalence curve for the random forest and k-nearest neighbors’ algorithms, trained using the *CBC dataset*. The points on the curves indicate the prevalence in the dataset.

**Suppl. Figure 13.**
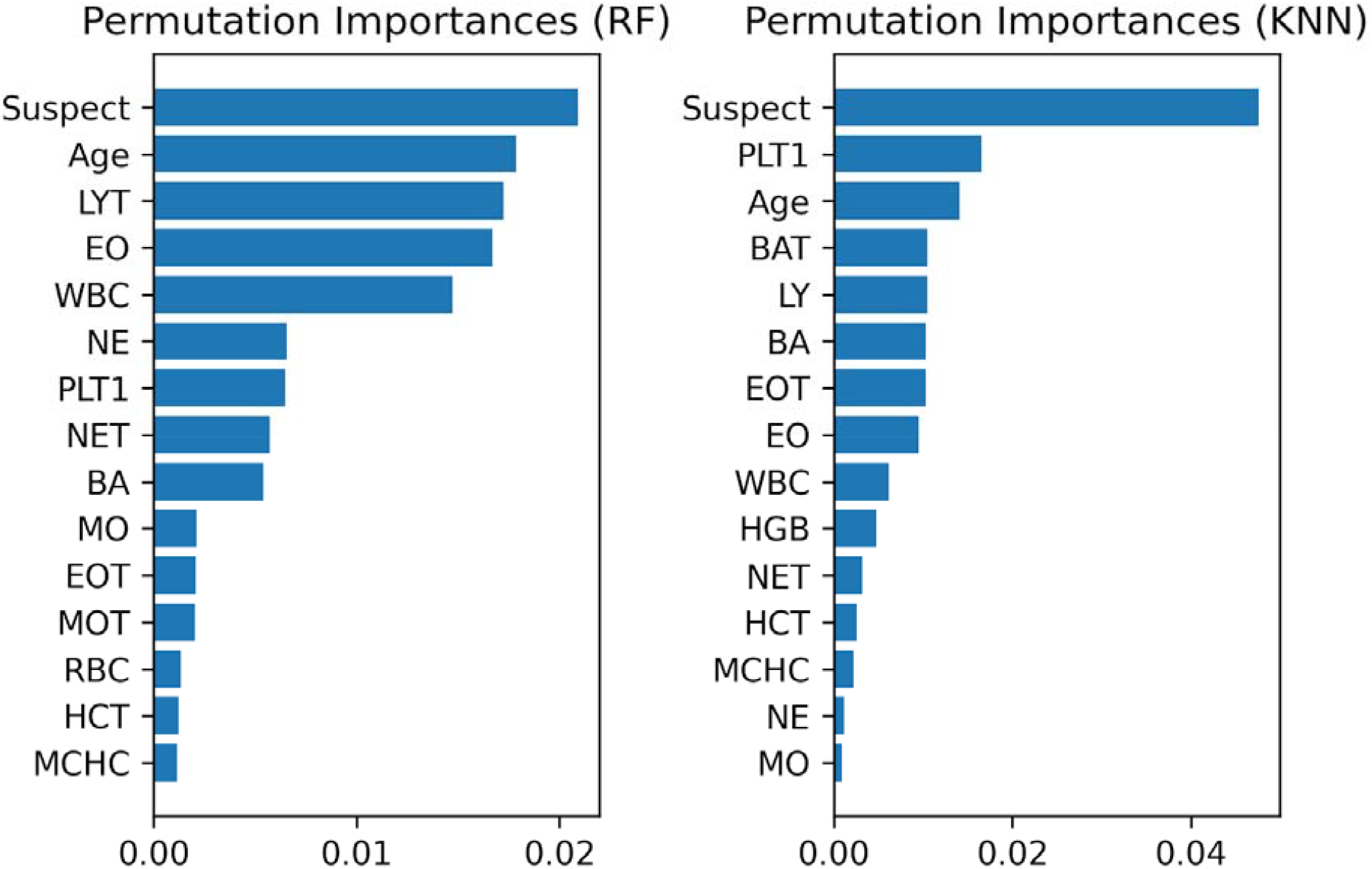
Feature importances for Random Forest and k-Nearest Neighbors, trained on the *CBC dataset*.

### Identification of the Uncertain Cases

In the ground truthing process, we identified 165 uncertain cases for which we combined the results of the rRT-PCR test together with the radiologic gold standard. The uncertain cases were identified through two different methods: either patients who turned out to be positive within 72 hours after a first negative test and were admitted as inpatients despite this test result; or patients who, despite having had a negative test, had a hematochemical profile that was more similar to positive patients. For this purpose we used the k-means clustering algorithm (k = 2) based on a set of COVID-19 characteristic biomarkers (AST, lymphocytes, calcium, LDH, CRP, WBC, XDP, fibrinogen) ^20,21^.

### Implementation of the Internal-External Validation

The internal-external validation was performed based on the *IOG dataset*, using a bootstrap-based procedure. The goal of this procedure was to evaluate the ability of the developed models to generalize to new settings when provided with a limited amount of new data.

First, we generated 100 random, 50/50 train-test splits of the *IOG dataset*, then for each of these splits: first, the train set of the *IOG dataset* was oversampled using the SMOTE algorithm to obtain a sample of 1,624 synthetic instances; second, the oversampled train set was combined with the *COVID-specific* (respectively, *CBC*) dataset to obtain a combined training set encompassing 3,248 instances; third, the best models (obtained as described in the Methods and Results sections) were re-trained over the combined training set and evaluated on the test set. The average results over the 100 generated splits were reported.

### Hematochemical Analysis

The hematological analyses were performed on a Sysmex XE 2100 system (Sysmex, Japan) and the coagulation features were determined using the STAR Max analyzer (Stago Group, France); the biochemical parameters were measured on a Roche COBAS 6000 system (Roche Diagnostic, Basel, Switzerland) using Roche reagents, calibrators (Calibrator for automated systems [Cfas]/Cfas proteins), and control materials at two different levels (Precicontrol ClinChem Multi 1 and 2). All of the methods for the enzyme activity measurements were standardized to IFCC reference measurement procedures. The point of care (POC) measurements and the hemogas analysis were undertaken using Rapidpoint 500 (Siemens Healthcare).

#### List of abbreviations

ALP: Alkaline Phosphatase
ALT: Alanine Aminotransferase
ANGPOC: Anion gap
AST: Aspartate Aminotransferase
AUC: Area under the curve
BA: Basophils count (%)
BAT: Basophils count
BEEPOC: Actual Base Excess
BEPOC: Base Excess
BICPOC: Bicarbonates
BILD: Direct Bilirubin
BILIN: Indirect Bilirubin
BILT: Total Bilirubin
BISPOC: Standard Calculated Bicarbonates
BO2POC: Bound O2 Maximum Concentration
CA: Calcium
CAPOC: Ionized Calcium (POC)
CASPOC: Standard Ionized Calcium(POC)
CBC: complete blood count
CK: Creatine kinase
CLPOC: Chloride (POC)
CO2POC: Carbonic Anhydride (pCO2)
CREA: Creatinine
CRP: C-reactive Protein
CT: computed tomography
CTOPOC: Total Oxygen
ED: emergency department
EO: Eosinophils count (%)
EOT: Eosinophils count
FCOPOC: Carboxyhemoglobin
FG: Fibrinogen
FIOPOC: Inspired Oxygen Fraction
FO2POC: Oxyhemoglobin / Total Hemoglobin
GGT: Gamma Glutamyltransferase
GLU: Glucose
GLUEM: Glucose Blood Gas
O
HCT: Hematocrit
HCTPOC: Hematocrit (POC)
HGB: Hemoglobin
HHBPOC: Deoxyhemoglobin
IL6: Interleukin 6
IOG: Istituto Ortopedico Galeazzi
K: Potassium
KNN: k-nearest neighbors
KPOC: Potassium (POC)
LATPOC: Lactate (POC)
LDH: Lactate Dehydrogenase
LR: logistic regression
LY: Lymphocytes count (%)
LYT: Lymphocytes count
MCH: Mean Corpuscolar Hemoglobin
MCHC: Mean Corpuscolar Hemoglobin Concentration
MCV: Average Globular Volume
METPOC: Methemoglobin
ML: machine learning
MO: Monocytes count (%)
MOT: Monocytes count
MPV: Average Platelet Volume
NA: Sodium
NAPOC: Sodium (POC)
NB: Naive Bayes
NE: Neutrophils count (%)
NET: Neutrophils count
NPV: Negative predictive Value
OFIPOC: Inspired O2 / O2 ratio
OSR: Ospedale San Raffaele
PCR: Polymerase Chain Reaction
PHPOC: pH
PLT: Platelets PO2POC Oxygen (pO2)
PPTR: Activated partial thromboplastin time (R)
PPV: positive predictive value
PROBNP: NT-proB-type Natriuretic Peptide
PTINR: Prothrombin Time (INR)
rRT-PCR: reverse transcription polymerase chain reaction
RBC: Red Blood Cells
RDW: Erythrocyte distribution width
RF: random forest
ROC: receiver operating characteristic
RT-PCR: reverse transcriptase–PCR
SO2POC: O2 Saturation
SVM: support vector machine
THBPOC: Total Oxyhemoglobin
TROPOT: Troponin T
UREA: Urea
WBC: White blood cells
XDP: D-Dimer

## References

1. Oran DP, Topol EJ. Prevalence of Asymptomatic SARS-CoV-2 Infection: A Narrative Review. [Published online June 3, 2020]. Ann intern med. doi:10.7326/M20-3012

2. Vogels CBF, Brito AF, Wyllie AL, Fauver JR, Ott IM, Kalinich CC, et al. Analytical sensitivity and efficiency comparisons of SARS-CoV-2 RT–qPCR primer–probe sets. [Published online July 10, 2020]. Nat Microbiol. doi:10.1038/s41564-020-0761-6

3. Lippi G, Simundic A-M, Plebani M. Potential preanalytical and analytical vulnerabilities in the laboratory diagnosis of coronavirus disease 2019 (COVID-19). Clin Chem Lab Med. 2020;58:1070–6

4. Woloshin S, Patel N, Kesselheim AS. False Negative Tests for SARS-CoV-2 Infection — Challenges and Implications. N Engl J Med. 2020;383:e38. doi: 10.1056/NEJMp2015897.

5. Wynants L, Van Calster B, Collins GS, Riley RD, Heinze G, Schuit E, et al. Prediction models for diagnosis and prognosis of covid-19: Systematic review and critical appraisal. BMJ. 2020; 369:m1328. doi: 10.1136/bmj.m1328

6. Li L, Qin L, Xu Z, Yin Y, Wang X, Kong B, et al. Artificial Intelligence Distinguishes COVID-19 from Community Acquired Pneumonia on Chest CT. [Published online April 3, 2020]. Radiology.doi:10.1148/radiol.2020200905

7. Gozes O, Frid-Adar M, Greenspan H, Browning PD, Zhang H, Ji W, et al. Rapid AI Development Cycle for the Coronavirus (COVID-19) Pandemic: Initial Results for Automated Detection & Patient Monitoring using Deep Learning CT Image Analysis.[Published online March 24, 2020]. arXiv Prepr arXiv http://arxiv.org/abs/2003.05037

8. Ozturk T, Talo M, Yildirim EA, Baloglu UB, Yildirim O, Rajendra Acharya U. Automated detection of COVID-19 cases using deep neural networks with X-ray images. Computers in Biology and Medicine 121 (2020) 103792. doi:10.1016/j.compbiomed.2020.103792

9. Mei X, Lee HC, Diao K, Huang M, Lin B, Liu C, et al. Artificial intelligence–enabled rapid diagnosis of patients with COVID-19. Nat Med 26, 1224–1228 (2020).

10. Weinstock MB, Echenique A, Russell JW, Leib A, Miller J, Cohen DJ, et al. Chest X-Ray Findings in 636 Ambulatory Patients with COVID-19 Presenting to an Urgent Care Center: A Normal Chest X-Ray Is no Guarantee. [Published online May, 2020]. JUCM. https://www.jucm.com/documents/jucm-covid-19-studyepub-april-2020.pdf/. Accessed August 17, 2020

11. Fan BE, Chong VCL, Chan SSW, Lim GH, Tan GB, Mucheli SS et al. Hematologic parameters in patients with COVID-19 infection. Am J Hematol. 2020;95:E131–E134.

12. Ferrari D, Motta A, Strollo M, Banfi G, Locatelli M. Routine blood tests as a potential diagnostic tool for COVID-19. Clin Chem Lab Med. 2020;58:1095–9.

13. Formica V, Minieri M, Bernardini S, Ciotti M, D’Agostini C, Roselli M, et al. Complete blood count might help to identify subjects with high probability of testing positive to SARS-CoV-2. Clin Med (Lond). 2020;20:e114–e119.

14. Wu J, Zhang P, Zhang L, Meng W, Li J, Tong C, et al. Rapid and accurate identification of COVID-19 infection through machine learning based on clinical available blood test results. [Published online 2020].medRxiv. doi:10.1101/2020.04.02.20051136

15. Soares F. A novel specific artificial intelligence-based method to identify {COVID}-19 cases using simple blood exams. [Published online 2020]. medRxiv. https://www.medrxiv.org/content/10.1101/2020.04.10.20061036v2

16. Soltan AAS, Kouchaki S, Zhu T, Kiyasseh D, Taylor T, Hussain ZB, et al. Artificial intelligence driven assessment of routinely collected healthcare data is an effective screening test for COVID-19 in patients presenting to hospital. [Published online 2020]. medRxiv.doi:10.1101/2020.07.07.20148361

17. Kukar M, Gunčar G, Vovko T, Podnar S, Černelč P, Brvar M, et al. COVID-19 diagnosis by routine blood tests using machine learning. [Published online June 2020]. arXiv Prepr arXiv.http://arxiv.org/abs/2006.03476. Accessed August 17, 2020

18. Collins GS, Moons KGM. Reporting of artificial intelligence prediction models. Lancet. 2019;393:1577–9.

19. Brinati D, Campagner A, Ferrari D, Locatelli M, Banfi G, Cabitza F. Detection of COVID-19 Infection from Routine Blood Exams with Machine Learning: A Feasibility Study. J Med Syst. 2020;44:135.

20. Collins GS, Reitsma JB, Altman DG, Moons KGM. Transparent reporting of a multivariable prediction model for individual prognosis or diagnosis (TRIPOD): The TRIPOD Statement. Eur Urol. 2015;131:211–9.

21. Watson J, Whiting PF, Brush JE. Interpreting a covid-19 test result. [Published online May 12, 2020].BMJ. doi:10.1136/bmj.m1808

22. Zitek T. The appropriate use of testing for Covid-19. West J Emerg Med. 2020 Apr 13;21:470–2.

23. Fang Y, Zhang H, Xie J, Lin M, Ying L, Pang P, et al. Sensitivity of Chest CT for COVID-19: Comparison to RT-PCR. Radiology. 2020;296:E115–E117.

24. Liu J, Yu H, Zhang S. The indispensable role of chest CT in the detection of coronavirus disease 2019 (COVID-19). Eur J Nucl Med Mol Imaging. 2020;47:1638–9.

25. Bohn MK, Lippi G, Horvath A, Sethi S, Koch D, Ferrari M, et al. Molecular, serological, and biochemical diagnosis and monitoring of COVID-19: IFCC taskforce evaluation of the latest evidence. Clin Chem Lab Med. 2020 25;58:1037–52.

26. Jadhav A, Pramod D, Ramanathan K. Comparison of Performance of Data Imputation Methods for Numeric Dataset. Appl Artif Intell. 2019;10:913–33

27. Guyon I, Weston J, Barnhill S, Vapnik V. Gene selection for cancer classification using support vector machines. Mach Learn. 2002;46:389–422.

28. Caruana R, Karampatziakis N, Yessenalina, A. An empirical evaluation of supervised learning in high dimensions. Proceedings of the 25th ICML. 2008;96–103

29. Du M, Liu N, Hu X. Techniques for interpretable machine learning. Communications of the ACM. 2019;63:68–77.

30. Brier GW. Verification of Forecasts Expressed in Terms of Probability. Mon Weather Rev. 1950;78:1–3.

31. Campagner A, Cabitza F, Ciucci D. The three-way-in and three-way-out framework to treat and exploit ambiguity in data. 2020;119: 292–312.

32. Banerjee A, Ray S, Vorselaars B, Kitson J, Mamalakis M, Weeks S, et al. Use of Machine Learning and Artificial Intelligence to predict SARS-CoV-2 infection from Full Blood Counts in a population. [Published online June 16, 2020]. Int Immunopharmacol. doi:10.1016/j.intimp.2020.106705

33. Avila E, Kahmann A, Alho C, Dorn M. Hemogram data as a tool for decision-making in COVID-19 management: applications to resource scarcity scenarios. [Published online June 29, 2020]. PeerJ. doi:10.7717/peerj.9482

34. Joshi RP, Pejaver V, Hammarlund NE, Sung H, Lee SK, Furmanchuk A, et al. A predictive tool for identification of SARS-CoV-2 PCR-negative emergency department patients using routine test results. J Clin Virol. 2020;129:104502. doi: 10.1016/j.jcv.2020.104502.

35. Yang HS, Vasovic L V, Steel P, Chadburn A, Hou Y, Racine-Brzostek SE, et al. Routine laboratory blood tests predict SARS-CoV-2 infection using machine learning. Clin Chem. [Published online August 21, 2020]. doi:10.1093/clinchem/hvaa200

36. Cabitza F, Campagner A, Ciucci D, Seveso A. Programmed Inefficiencies in DSS-Supported Human Decision Making. In: Lecture Notes in Computer Science (Including Subseries Lecture Notes in Artificial Intelligence and Lecture Notes in Bioinformatics). 2019. doi:10.1007/978-3-030-26773-5_18

37. Rodriguez-Morales AJ, Cardona-Ospina JA, Gutiérrez-Ocampo E, Villamizar-Peña R, Holguin-Rivera Y, Escalera-Antezana JP, et al. Clinical, laboratory and imaging features of COVID-19: A systematic review and meta-analysis. Travel Med Infect Dis. 2020;34:101623. doi:10.1016/j.tmaid.2020.101623

38. Zhang ZL, Hou YL, Li DT, Li FZ. Laboratory findings of COVID-19: a systematic review and meta-analysis. [Published online May 23, 2020] Scand J Clin Lab Invest.;1–7. doi: 10.1080/00365513.2020.1768587

39. Connors JM, Levy JH. COVID-19 and its implications for thrombosis and anticoagulation. Blood. 2020;135:2033–40.

40. Rabanser S, Günnemann S, Lipton ZC. Failing Loudly: An Empirical Study of Methods for Detecting Dataset Shift. 2018;(NeurIPS). http://arxiv.org/abs/1810.11953

41. Augenblick N, Kolstad JT, Obermeyer Z, Wang A. Group Testing in a Pandemic: The Role of Frequent Testing, Correlated Risk, and Machine Learning. Natl Bur Econ Res. 2020; http://www.nber.org/papers/w27457.pdf

42. Larremore DB, Wilder B, Lester E, Shehata S, Burke JM, Hay JA, et al. Test sensitivity is secondary to frequency and turnaround time for COVID-19 surveillance. [Published online 2020]. medRxiv. doi:10.1101/2020.06.22.20136309

43. Song JY, Yun JG, Noh JY, Cheong HJ, Kim WJ. Covid-19 in South Korea - Challenges of subclinical manifestations. N Engl J Med. 2020; 382:1858–9

44. Service R. Fast, cheap tests could enable safer reopening. Science. 2020;369:608–9.

